# Efficacy and Safety of Acupuncture for Acute Gouty Arthritis: A Systematic Review and Network Meta-Analysis

**DOI:** 10.1101/2024.09.21.24314132

**Authors:** Yuxiao Cai, Yifan Liu, Qinglong Kang, Shiji Zhu, Qianqian Wang, Chunmei Sun, Yin Gu

## Abstract

Acute gouty arthritis (AGA) severely affects the quality of life of patients due to its persistent, recurrent, and debilitating characteristics. Acupuncture holds a pivotal role as a classical alternative therapy in managing gouty arthritis. This study aimed to assess the clinical effectiveness of various acupuncture methods used in conjunction with conventional medications for treating acute gouty arthritis. We conducted a search of seven English and Chinese databases up to February 10, 2024, and analyzed the data using R software version 4.4.0.The study included a total of 37 randomized controlled trials(RCT), encompassing 2,801 patients. The network meta-analysis (NMA) results are as follows: Fire acupuncture demonstrated the highest effectiveness in improving cure rates. Acupuncture combined with conventional medication was found to be the most effective for reducing VAS scores. Additionally, Acupuncture combined with conventional medication proved most effective in lowering uric acid levels. For reducing ESR, the combination of blood-letting therapy and conventional drugs showed the greatest efficacy. No statistically significant differences were found among the different interventions regarding the reduction of TCMSS and CRP levels. In conclusion, fire acupuncture showed the greatest overall effectiveness. Acupuncture with conventional medication best reduced VAS scores and uric acid levels, while blood-letting therapy combined with conventional drugs was most effective in lowering ESR.

## 1 Introduction

Gouty arthritis (GA) is an inflammatory condition resulting from the deposition of urate crystals in various tissues, including the joint capsule, bursa, cartilage, and surrounding structures. It is prevalent in men and can be prevented by oral uric acid-lowering drugs, along with lifestyle modifications such as weight loss and adhering to a low-purine diet, to prevent gout flares. Acute gouty arthritis typically manifests with sudden onset at night, characterized by severe pain in the affected joints, accompanied by redness, swelling, heat, tenderness to touch, weakness, fever, headache, and other symptoms^1^. Currently, there is no definitive cure for gouty arthritis. The American College of Rheumatology (ACR) and the European League Against Rheumatism (EULAR) advise managing gouty arthritis with medications such as non-steroidal anti-inflammatory drugs (NSAIDs), glucocorticoids, and colchicine. However, many side effects (e.g., gastrointestinal adverse reactions, jaundice, proteinuria, etc.) are produced by these drugs in the course of treatment^2–4^. Acupuncture is a complementary and alternative therapy rooted in the meridian theory of traditional Chinese medicine. It has been extensively used for the treatment of conditions such as knee osteoarthritis, ankylosing spondylitis, and various other forms of arthritis, often demonstrating encouraging clinical results ^5–7^. Acupuncture is commonly used in conjunction with conventional drugs for the treatment of gouty arthritis. Previous studies have confirmed that acupuncture combined with conventional drugs is more effective than conventional drugs alone^5^. However, various types of acupuncture such as electroacupuncture, moxibustion, and fire acupuncture exist, yet direct comparisons of their efficacy remain scarce. Therefore, the selection of appropriate acupuncture treatments in clinical practice remains contentious. Network meta-analysis (NMA) advances beyond conventional pairwise meta-analysis techniques. Based on the current study, NMA enables direct and indirect comparisons of various acupuncture therapies. By synthesizing and evaluating various comparisons, this approach ranks the relative effectiveness of different acupuncture therapies. Consequently, this study utilized NMA to assess the relative efficacy of multiple acupuncture methods for treating gouty arthritis, with the goal of identifying the most effective treatment strategies for clinical practice.

## 2 Materials and methods

### 2.1 Register

The network meta-analysis was performed in line with the PRISMA-NMA guidelines. The research has been formally registered with PROSPERO, and details can be accessed via PROSPERO under registration number CRD42021278233.

### 2.2 Criteria for Inclusion and Exclusion

#### 2.2.1 Study Design

This review incorporated randomized controlled trials (RCTs) published in both English and Chinese languages.

#### 2.2.2 Participant Criteria

Participants with acute gouty arthritis were required to meet specific diagnostic criteria, such as the 2015 Gout Classification Criteria established by the American College of Rheumatology and the European League Against Rheumatism, without regard to their age or sex ^8^.

#### 2.2.3 Treatment Protocols

The intervention for patients in the experimental group involved a regimen combining various acupuncture therapies with standard medications. These therapies included conventional acupuncture, Patients in the treatment group received a combination of acupuncture-related therapies along with conventional drugs. The therapeutic approaches encompassed conventional acupuncture, warming acupuncture, electroacupuncture, fire acupuncture, blood-letting, moxibustion, buried acupuncture therapy, and acupoint injections. In contrast, patients in the control group received treatment with either conventional medications alone or combined with acupuncture-related therapies. Consistency in the use of different conventional drugs, whether administered alone or in combination, was maintained across both groups in the study.

#### 2.2.4 Outcome Measures

The primary outcome measure was the cure rate, characterized by the resolution of clinical symptoms and normalization of serum marker criteria. The secondary outcome indicators encompassed the Visual Analogue Scale (VAS), serological markers such as C-reactive protein (CRP) and erythrocyte sedimentation rate (ESR), blood uric acid (UA), and adverse reactions (AEs).

#### 2.2.5 Exclusion Criteria

(1). Lack of clear diagnosis criteria; (2). Absence of defined outcome indicators; (3) Inclusion of multiple Traditional Chinese Medicine (TCM) therapies, including cupping, traditional Chinese herbs, and combinations such as acupuncture with moxibustion and electroacupuncture; (4). Duplicate publication of studies; (5). Incomplete data despite attempts to contact the authors.

### 2.3 Literature Search Strategy

We searched PubMed, EMBASE, Web of Science, Cochrane Library, China National Knowledge Infrastructure (CNKI), Wan Fang, and VIP databases for randomized controlled trials (RCTs) investigating acupuncture combined with conventional drugs for the treatment of acute gouty arthritis. We utilized a comprehensive set of search terms including acupuncture, electroacupuncture, warm acupuncture, fire acupuncture, blood-letting therapy, moxibustion, acupoint catgut embedding, acupoint injection, acupuncture-knife therapy, and AGA in both Chinese and English languages. The PubMed database search strategy is outlined in Table 1.

**TABLE 1.** PubMed database retrieval strate.

| NUMBER | SEARCH TERMS |
| --- | --- |
| #1 | Gouty Arthritis[MESH] |
| #2 | Gouty Arthritis[Title/Abstract] |
| #3 | Gouty Arthritis[Title/Abstract] |
| #4 | gout*[Title/Abstract] |
| #5 | Acupuncture[MESH] |
| #6 | Acupuncture[Title/Abstract] |
| #7 | Ear Acupuncture[MESH] |
| #8 | Ear Acupuncture[Title/Abstract] |
| #9 | electro-acupuncture[Title/Abstract] |
| #10 | electroacupuncture[Title/Abstract] |
| #11 | Moxabustion[Title/Abstract] |
| #12 | warm acupuncture[Title/Abstract] |
| #13 | fire acupuncture [Title/Abstract] |
| #14 | blood-letting therapy[Title/Abstract] |
| #15 | Acupotomy[Title/Abstract] |
| #16 | acupoint catgut embedding[Title/Abstract]) |
| #17 | acupoint injection[Title/Abstract] |
| #18 | #1 OR #2 OR #3 OR #4 |
| #19 | #5 OR #6 OR #7 OR #8 OR #9 OR #10 OR #11 OR #12 OR #13 OR #14 OR #15 OR #16 OR #17 |
| #20 | #18 AND #19 |

### 2.4 Literature Review and Data Extraction

Duplicates were removed using EndNote X9 software, followed by an initial screening of each study based on its title and abstract. Subsequently, full texts were screened to exclude studies not meeting the inclusion criteria. Data extraction was conducted independently by two researchers (Cai and Liu) according to predefined criteria. Disagreements were resolved by a third researcher (Zhu) to reach a final decision. In case of any disagreement, the third researcher (Zhu) will arbitrate and make the final decision. Information extraction involves capturing details such as title, authors, publication year and month, diagnostic criteria used, sample sizes, details of interventions in both treatment and control groups including doses, treatment procedures, and outcome indicators.

### 2.5 Evaluation of the Risk of Bias

Two independent researchers (Cai and Liu) will assess quality using the RCT risk of bias assessment tool from the Cochrane Handbook of Systematic Reviews version 2.0.Any discrepancies between the two researchers will be resolved with assistance from a third researcher (Zhu). Evaluation criteria included Randomization process、Deviations from intended interventions、Missing outcome data、Measurement of the outcome and Selection of the reported result.

### 2.6 Statistical Analysis

R 4.4.0 was employed to create an evidence network diagram illustrating the comparative relationships among interventions for each outcome indicator. Small sample effects and publication bias were assessed using comparison-correction funnel plots. Simultaneously, a network meta-analysis was conducted using R 4.4.0, implementing the Markov Chain Monte Carlo (MCMC) consistent model within a Bayesian framework. The simulation involved four chains with a total of 50,000 iterations—20,000 for annealing and 30,000 for sampling. The assumption of MCMC convergence was assessed using the Potential Scale Reduction Factor (PSRF), while the stability and consistency of the results were evaluated using the MCMC fitting model.

The mean difference (MD) was used for continuous data, and odds ratios were used for dichotomous data, with all results reported using 95% confidence intervals (CI).To rank the cumulative probability of each outcome measure for each intervention, the surface under the cumulative ranking (SUCRA) was utilized. A higher SUCRA value indicated a more effective treatment option.

## 3 Results

### 3.1 Literature Screen

Initially, 3,060 papers were retrieved for this study. After thorough screening and re-screening, 37 randomized controlled trials (RCTs) involving 2,801 patients were included^5, 9–44^. The literature screening process proceeded as follows: Figure1

**Figure 1.**
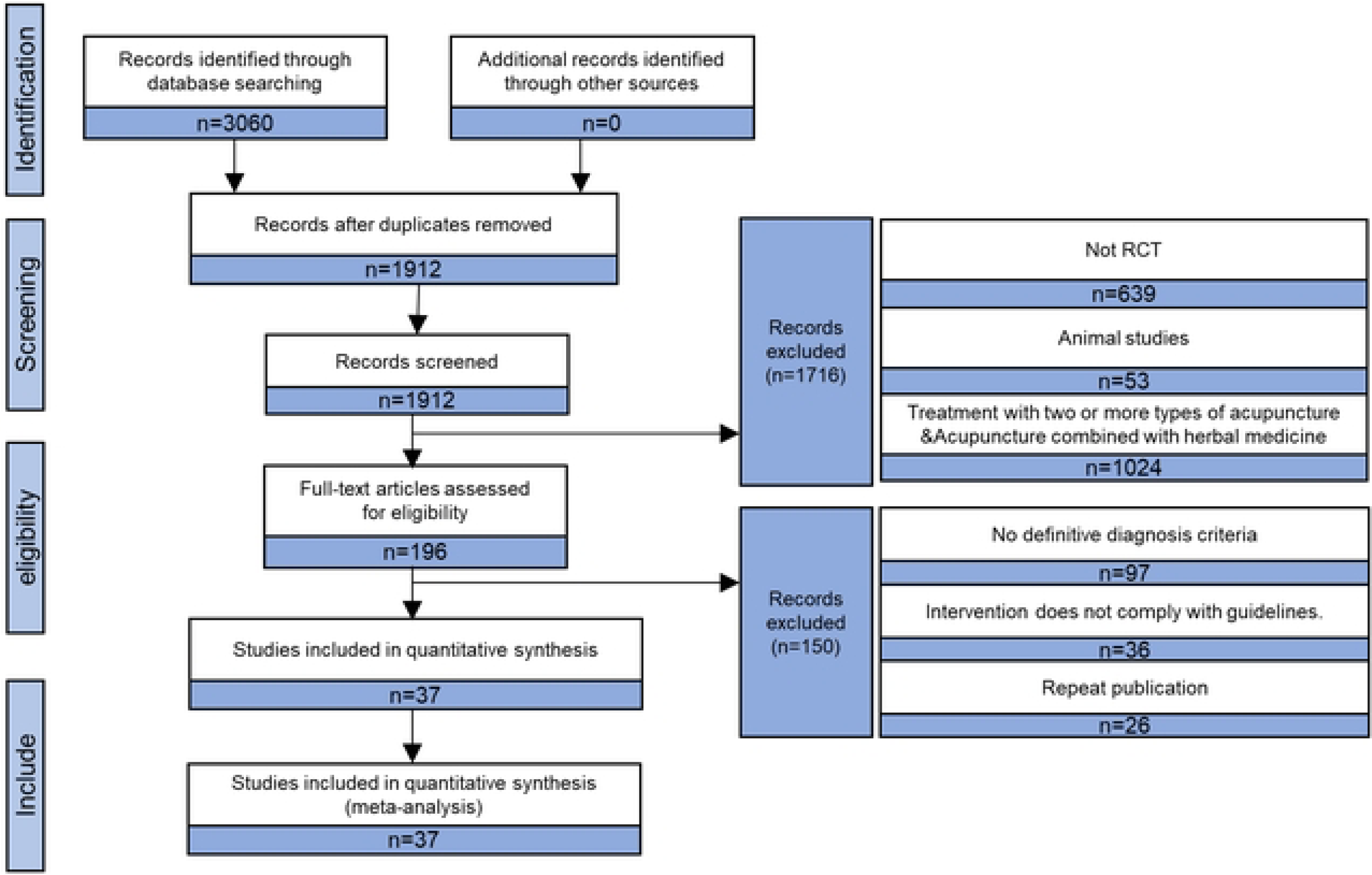
Flow diagram outlining the guideline selection proc.

### 3.2 Characteristics of the Included Studies

Among the 37 included studies, the distribution of therapies was as follows: 7 studies ^10, 20, 24, 27, 32, 34, 37^ reported acupuncture therapy; 6 studies ^9, 13, 31, 38, 39, 41^ reported acupuncture combined with conventional drugs, 2 studies ^19, 23^ reported acupotomy therapy; 2 studies^17, 21^ reported acupotomy combined with conventional drugs; 8 studies ^12, 18, 25, 26, 29, 36, 40, 43, 44^ reported blood-letting therapy; 3 studies ^28, 30, 36^ reported electroacupuncture combined with conventional drugs; 37 studies ^5, 9–44^,3 studies ^5, 16, 33^ reported electroacupuncture combined with conventional drugs; 3 studies ^14, 15, 35^ reported fire acupuncture; 2 studies ^22, 42^ reported warm acupuncture;1 study ^11^ reported warm acupuncture combined with conventional drugs. Among the included studies: 36 studies ^9–44^ were two-arm studies and 1 study ^5^ was a three-arm study.

The reported outcomes were distributed as follows:Visual Analog Scale (VAS) scores: Reported in 17 studies (5, 10, 11, 16, 19, 23, 27-29, 32-36, 38, 40, 41);Adverse Events (AEs): Reported in 10 studies (10, 11, 16, 22, 25, 27, 28, 32, 33, 38).Cure rates: Reported in 32 studies ^9–20, 22–24, 26–31, 34–44^; Traditional Chinese Medicine Symptom Scores (TCMSS): Reported in seven studies^11, 14, 31, 36, 38, 39, 42^; C-reactive protein (CRP): Reported in eight studies ^11, 28, 31–33, 36, 39, 43^; Uric acid (UA): Reported in 27 studies ^9–11, 13–15, 17–21, 23–25, 27–29, 31, 32, 34, 36, 38–40, 42–44^; Erythrocyte sedimentation rate (ESR): Reported in nine studies ^11, 13, 25, 28, 31, 32, 36, 39, 43^; Visual Analog Scale (VAS) scores: Reported in 17 studies ^5, 10, 11, 16, 19, 23, 27–29, 32–36, 38, 40, 41^; Adverse Events (AEs): Reported in 10 studies ^10, 11, 16, 22, 25, 27, 28, 32, 33, 38^. Tables 2 and 3 outline the key characteristics of the included studies and interventions, respectively.

**Table 2.**
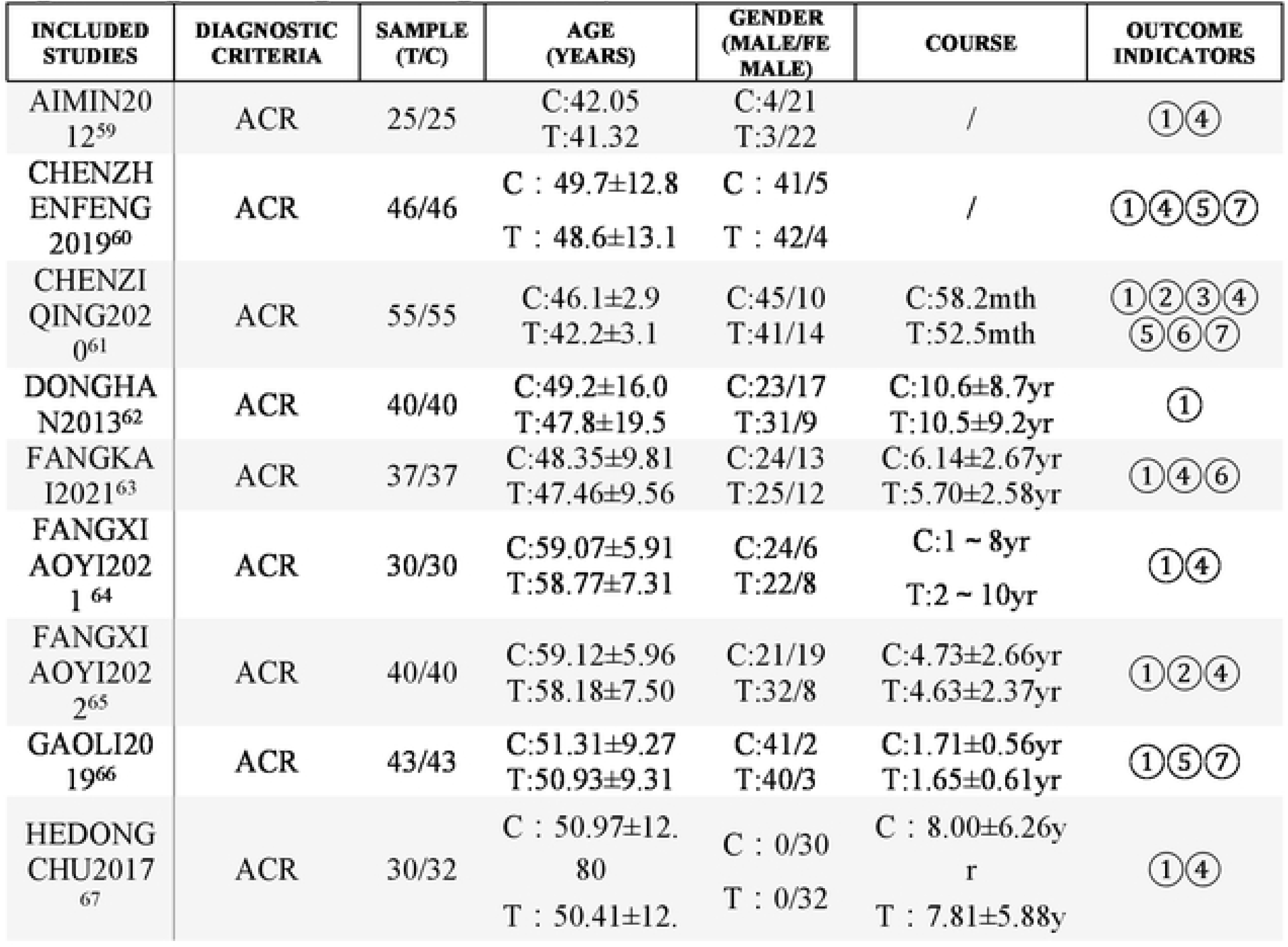
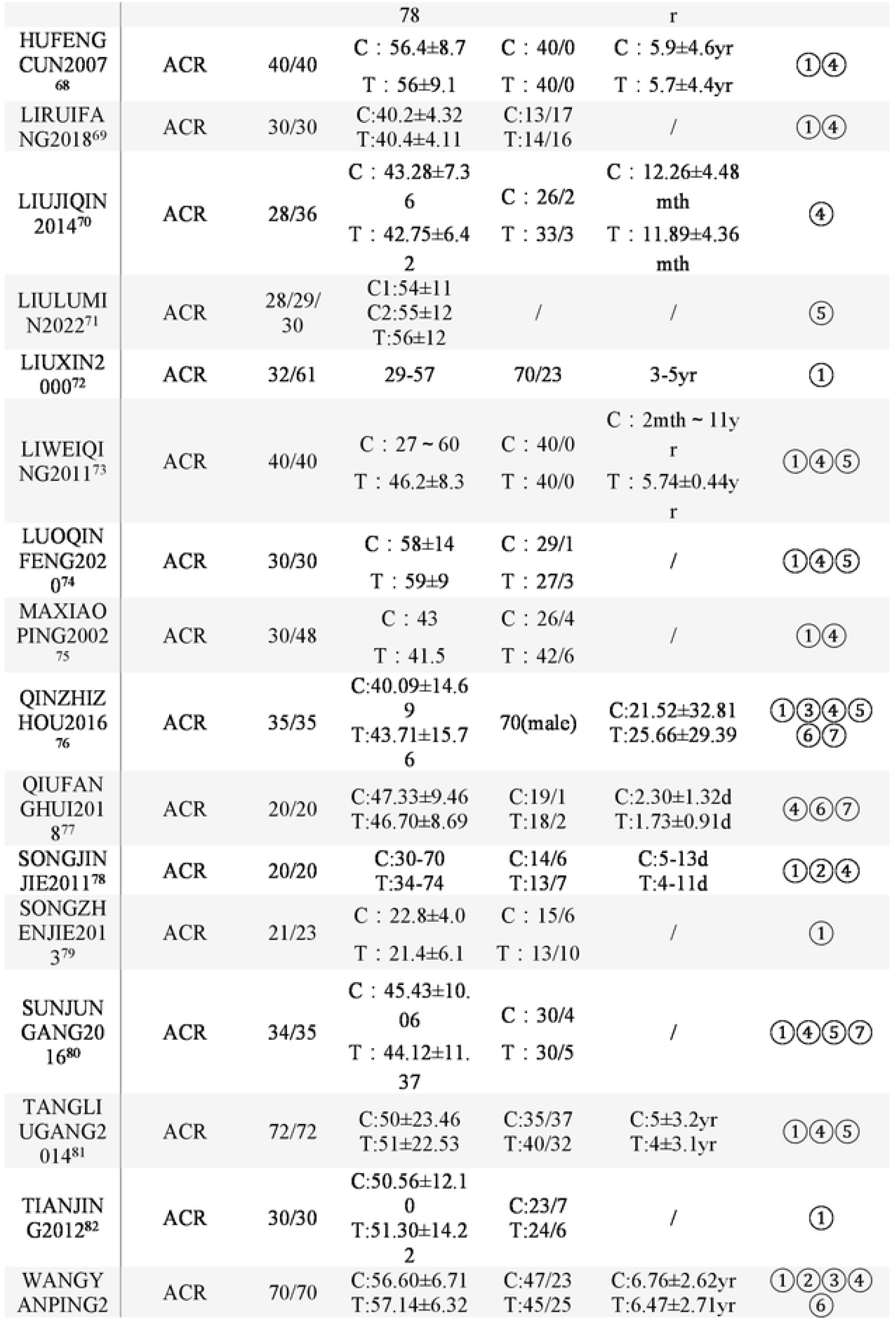
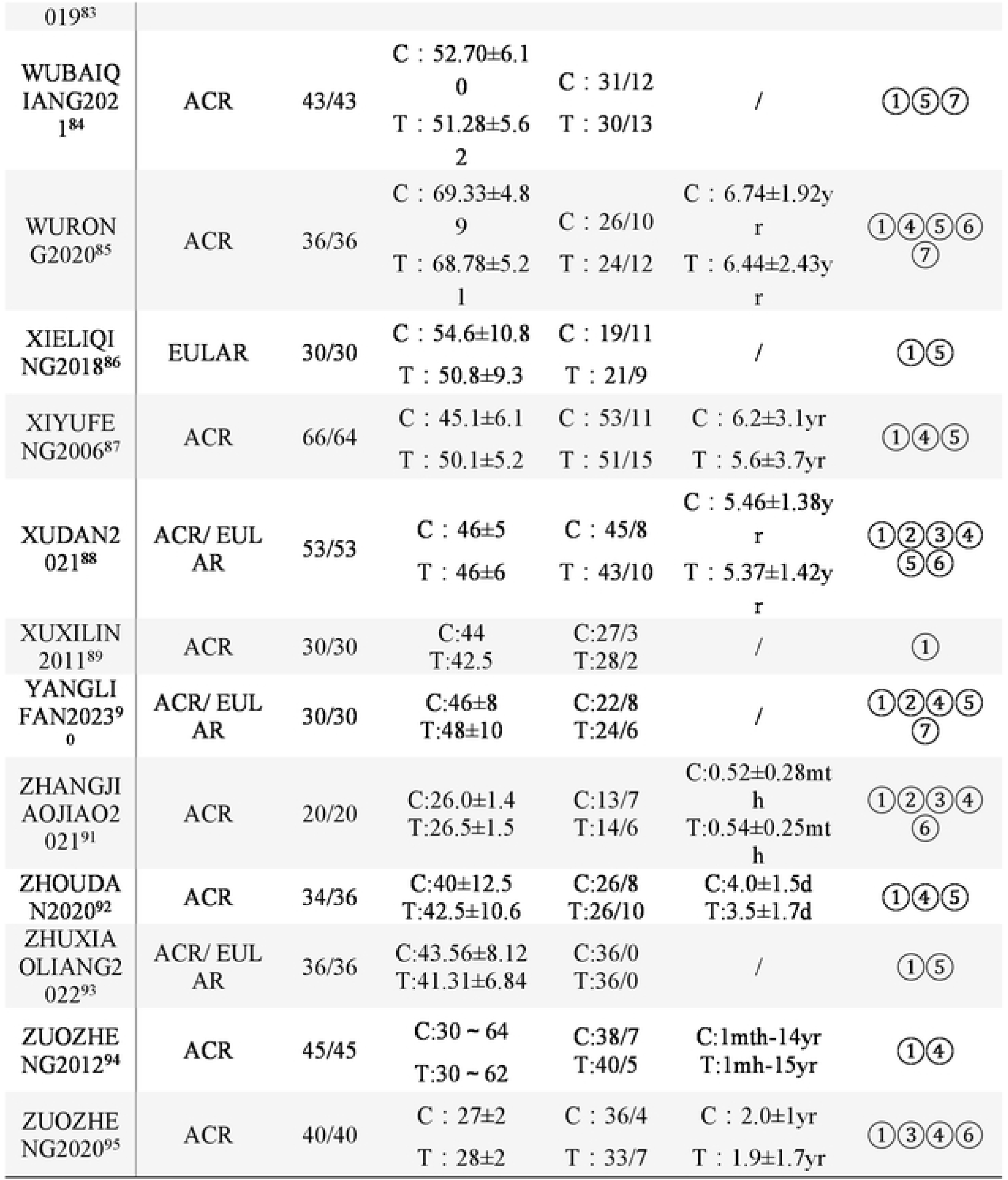
Characteristics of the included studies. (①Cute Rate; ②TCMSS; ③CRP; ④UA; ⑤VAS; ⑥ESR; ⑦Adverse)

**Table 3.**
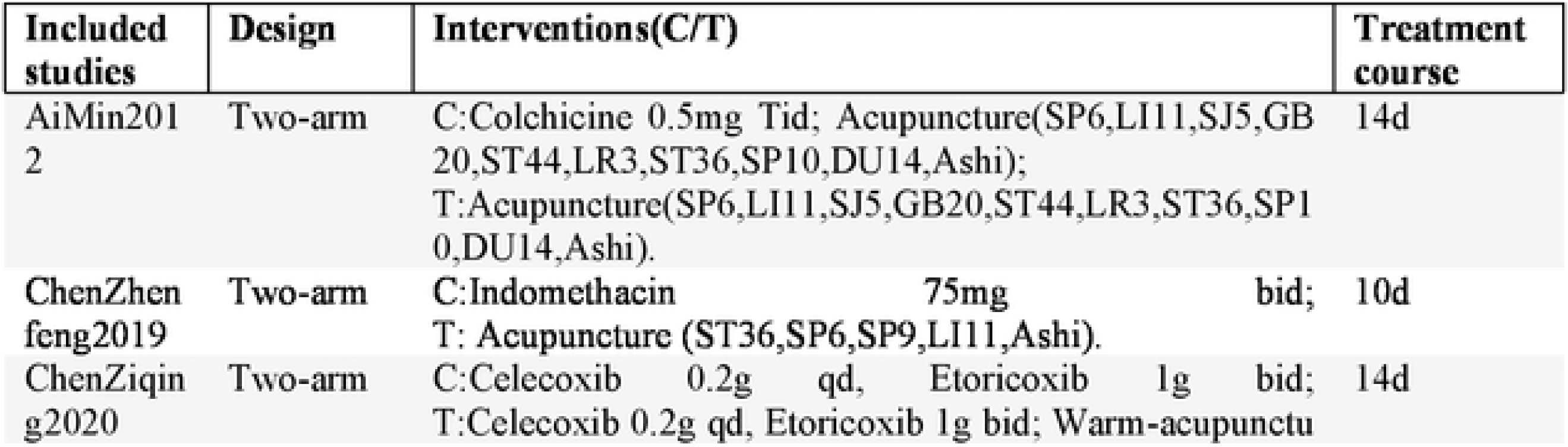
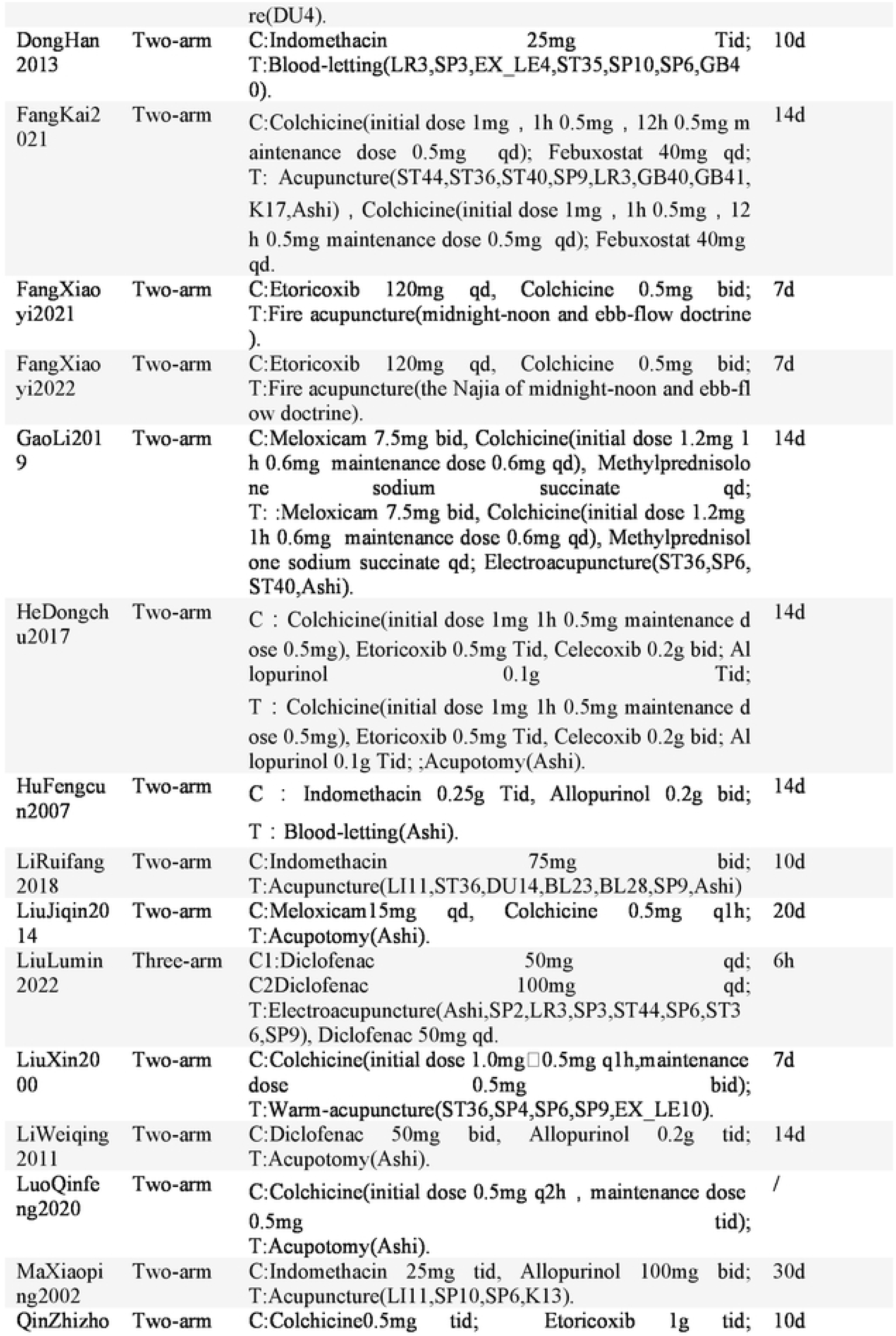
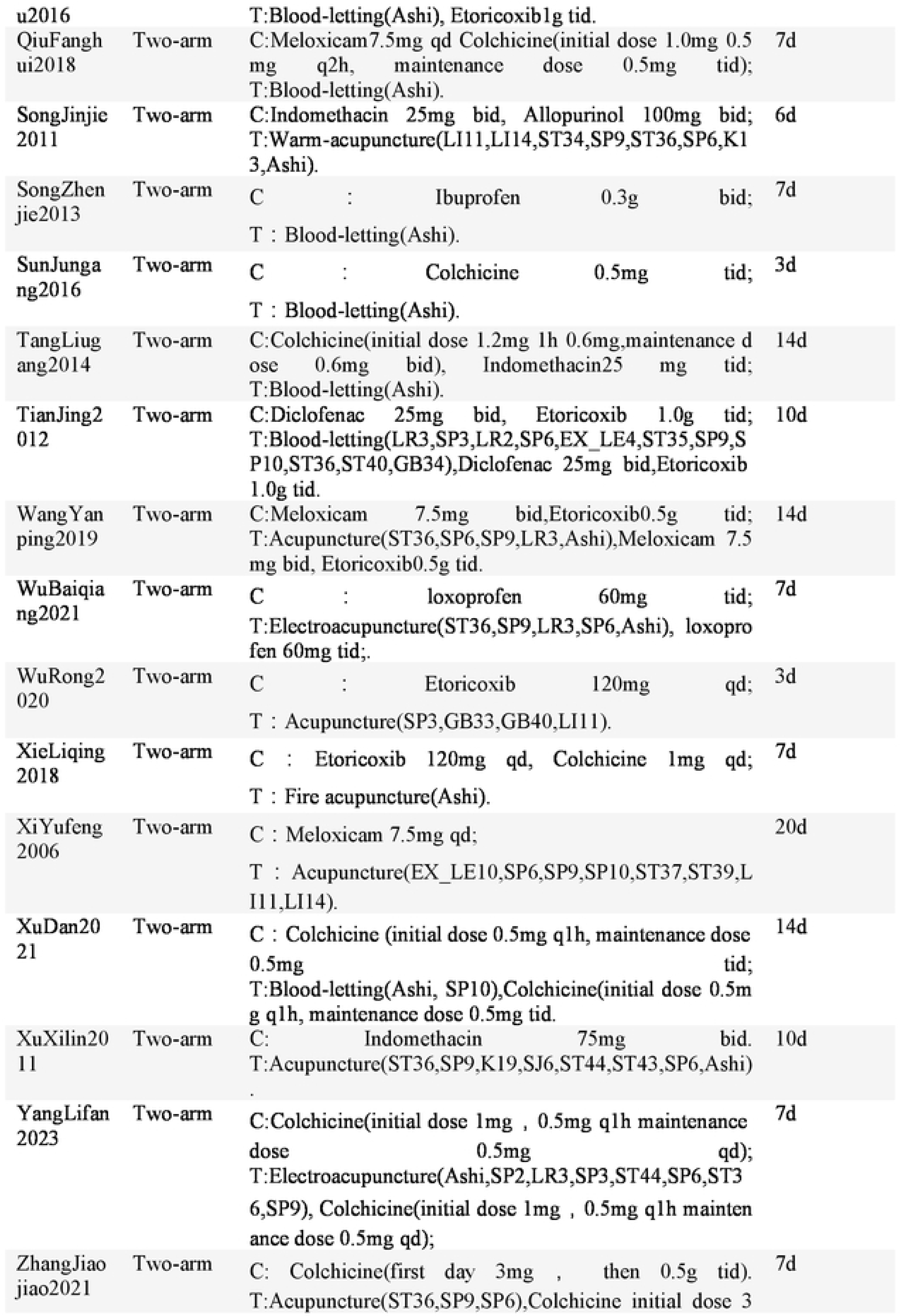
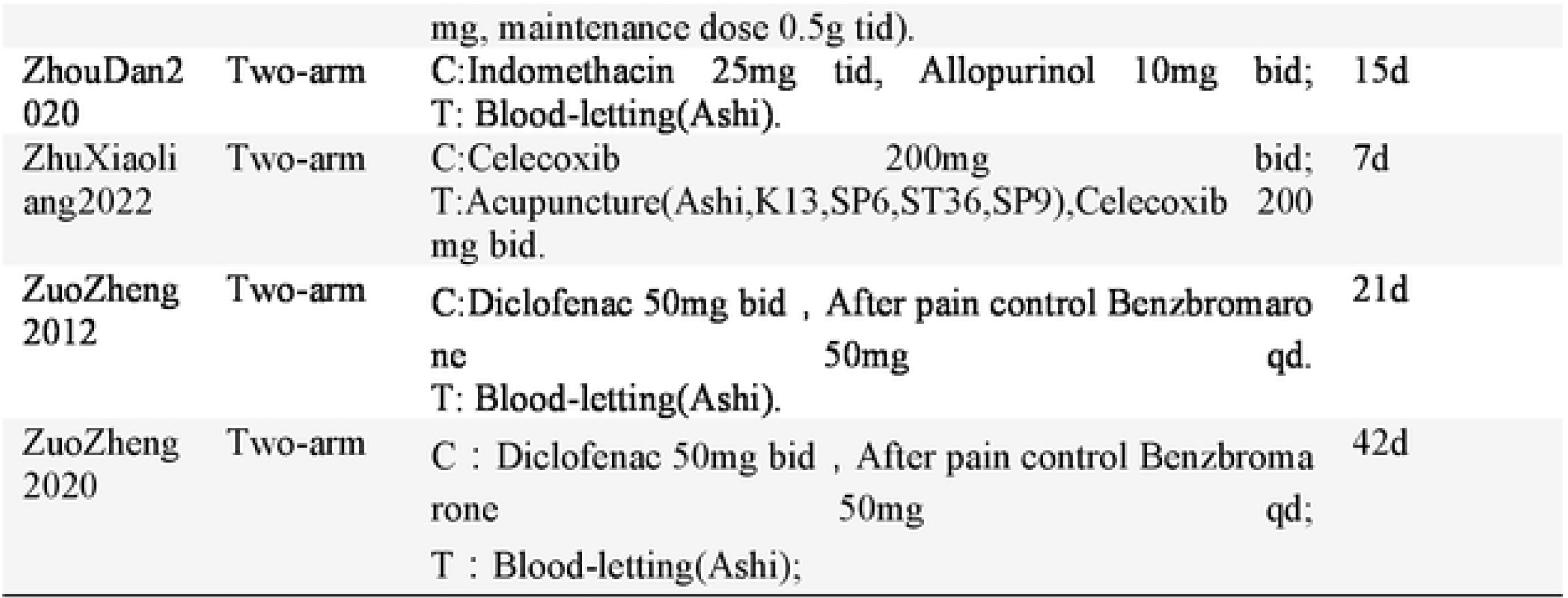
Characteristics of interventions of included studies.

**Table 4.**
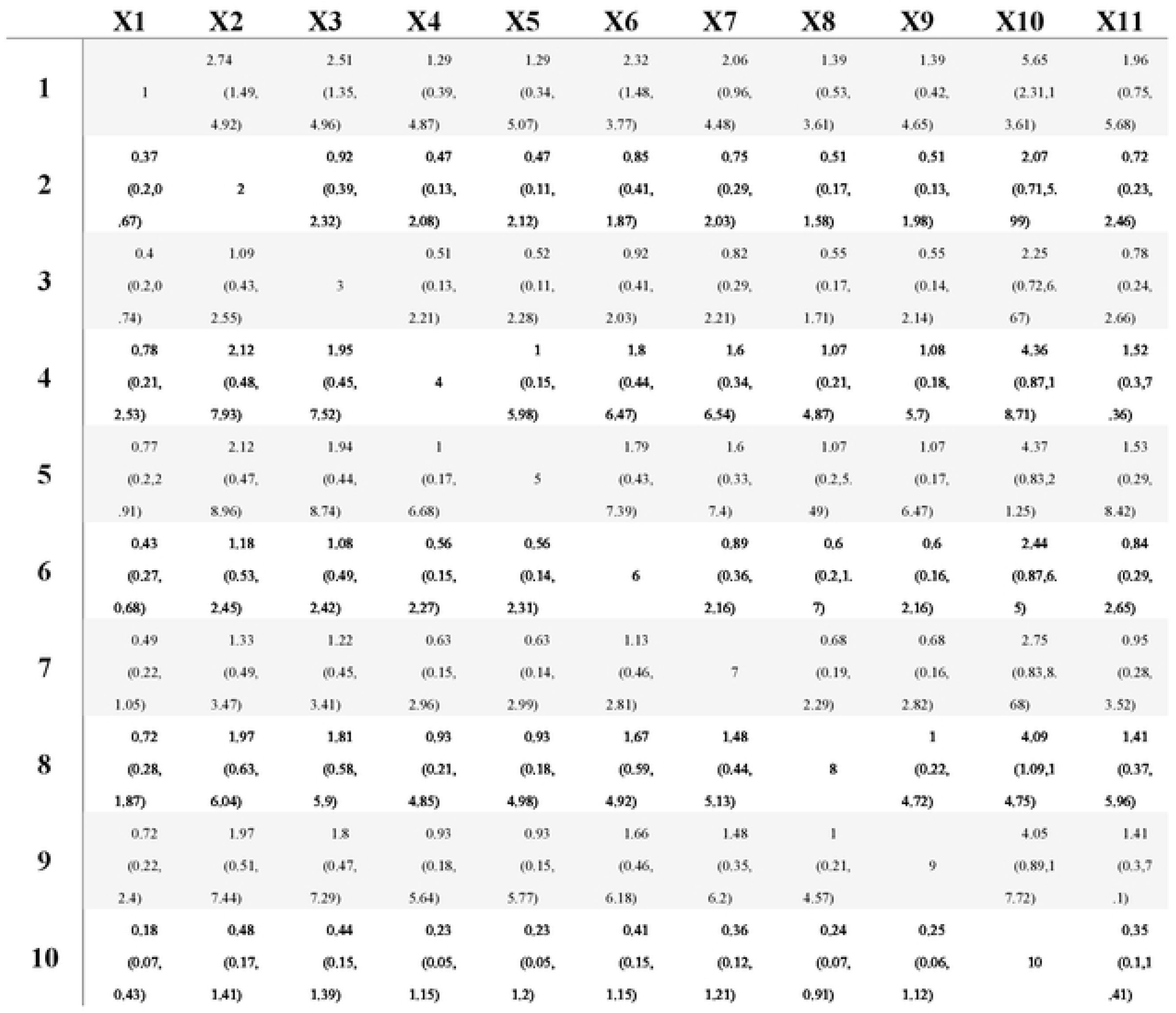
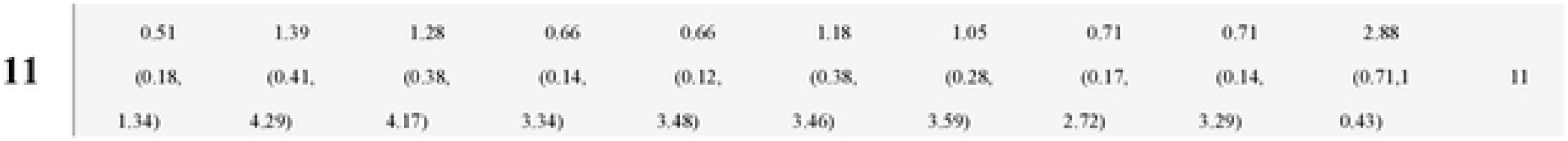
NMA results of the cure rate. (I Conventional drugs; 2 Acupuncture; 3 Acupuncture+ conventional drugs; 4 Acupotomy; 5 Acupotomy + conventional drugs; 6 Blood-letting; 7 Blood-letting therapy + conventional drugs; 8 Electroacupuncture + conventional drugs; 9 Warm acupuncture 9 + conventional drugs; 10 Fire acupuncture; 11 Warm acupuncture.)

**Table 5.** Probability ranking results of the cure rate. (1 Conventional drugs; 2 Acupuncture; 3 Acupuncture+ conventional drugs; 4 Acupotomy; *5* Acupotomy + conventional drugs; 6 Blood-letting; 7 Blood-letting therapy+ conventional drugs; 8 Electroacupuncture + conventional drugs;9 Warm acupuncture+ conventional drugs; 10 Fire acupuncture; 11 Warm acupuncture.)

|  | V1 | V2 | V3 | V4 | V5 | V6 | V7 | V8 | V9 | V10 | V11 |
| --- | --- | --- | --- | --- | --- | --- | --- | --- | --- | --- | --- |
| 1 | 0.000 | 0.000 | 0.000 | 0.000 | 0.000 | 0.001 | 0.017 | 0.097 | 0.284 | 0.392 | 0.209 |
| 9 | 0.019 | 0.050 | 0.048 | 0.053 | 0.063 | 0.085 | 0.118 | 0.140 | 0.131 | 0.127 | 0.167 |
| 10 | 0.778 | 0.119 | 0.046 | 0.025 | 0.015 | 0.009 | 0.005 | 0.002 | 0.001 | 0.000 | 0.000 |
| 11 | 0.040 | 0.106 | 0.098 | 0.098 | 0.107 | 0.128 | 0.135 | 0.120 | 0.082 | 0.050 | 0.035 |
| 2 | 0.045 | 0.252 | 0.221 | 0.172 | 0.128 | 0.088 | 0.054 | 0.027 | 0.010 | 0.003 | 0.000 |
| 3 | 0.038 | 0.174 | 0.188 | 0.176 | 0.152 | 0.121 | 0.081 | 0.047 | 0.019 | 0.004 | 0.001 |
| 4 | 0.021 | 0.050 | 0.043 | 0.047 | 0.056 | 0.072 | 0.101 | 0.128 | 0.127 | 0.135 | 0.220 |
| 5 | 0.024 | 0.053 | 0.046 | 0.048 | 0.055 | 0.072 | 0.098 | 0.121 | 0.118 | 0.129 | 0.235 |
| 6 | 0.010 | 0.082 | 0.159 | 0.209 | 0.218 | 0.165 | 0.099 | 0.043 | 0.013 | 0.002 | 0.000 |
| 7 | 0.020 | 0.089 | 0.113 | 0.125 | 0.143 | 0.159 | 0.149 | 0.107 | 0.058 | 0.026 | 0.010 |
| 8 | 0.007 | 0.026 | 0.037 | 0.047 | 0.062 | 0.098 | 0.143 | 0.168 | 0.157 | 0.131 | 0.123 |

### 3.3 Results of the Risk of Bias

Various methods were used to achieve random sequences and allocation concealment in the reviewed studies. Specifically, 19 studies utilized randomized number tables ^9–11, 13, 14, 16, 25–29, 33–39, 42^, one study employed lottery randomization ^12^, and single studies used random envelopes ^5^ and interval random methods (reference 43), respectively^43^. The remaining 15 studies^15, 17–24, 30–32, 40, 41, 44^ mentioned the use of randomization without specifying the method. In terms of blinding of investigators and subjects, neither blinding was reported due to intervention limitations. Regarding incomplete reporting, selective reporting, and other biases, all 37 studies included in the analysis provided complete data without selective reporting or other biases. The results of the risk of bias assessment are presented in Figure 2.

**Figure 2.**
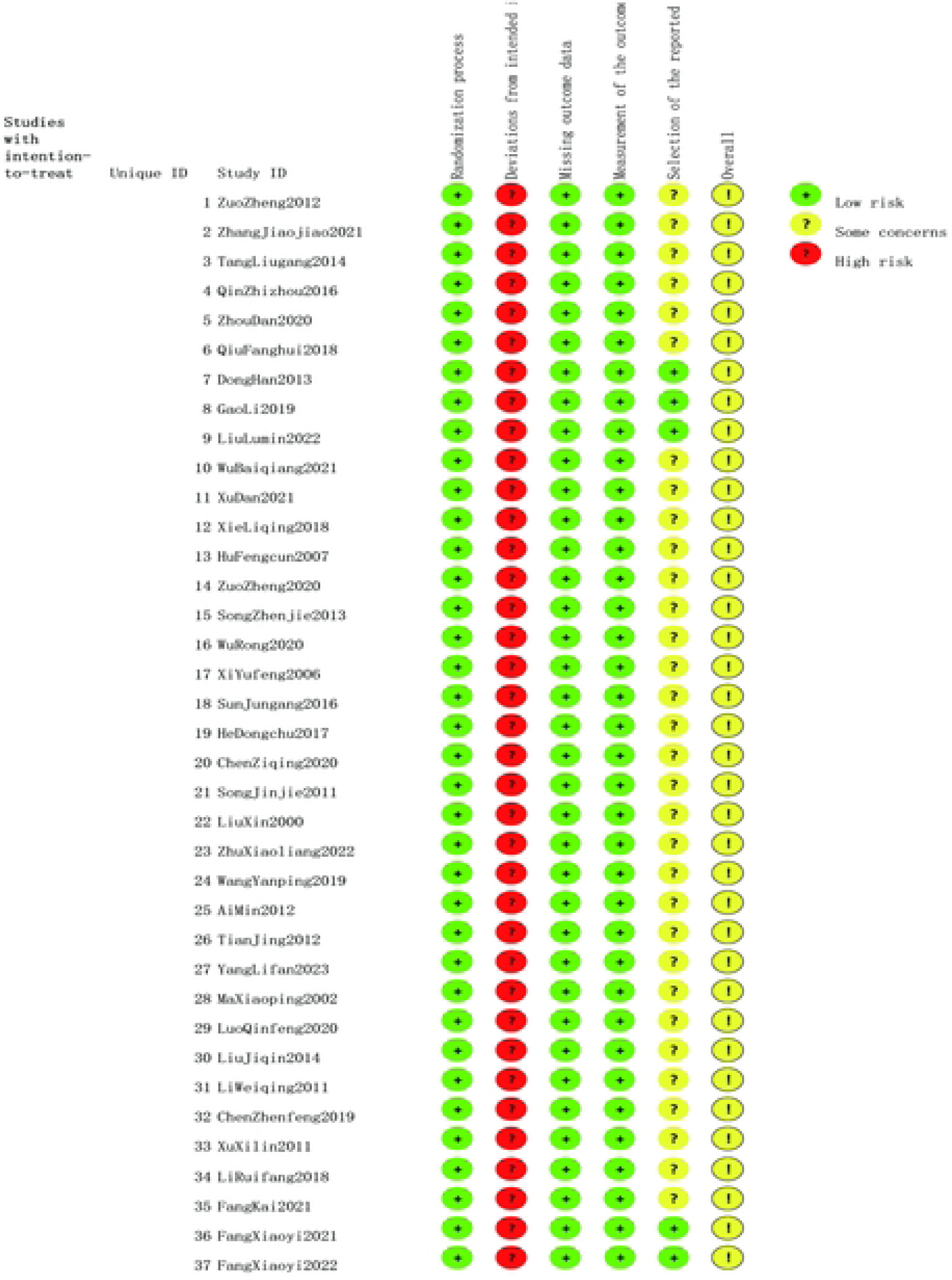
Results of the risk of bias evaluation.

### 3.4 Directly Compared Meta-Analysis Results

In terms of cure rate, acupuncture therapy, acupuncture combined with conventional drugs, blood-letting therapy, blood-letting therapy combined with conventional drugs, and fire acupuncture demonstrated superior efficacy compared to conventional drugs. There was no statistically significant difference observed between acupotomy, warm acupuncture therapy, electroacupuncture combined with conventional drugs, and conventional drugs. Other therapies have been studied in too few relevant studies to draw meaningful comparisons with conventional drugs. No relevant comparisons between different acupuncture treatments were conducted, as no cases were included where these treatments were directly compared with each other.

Acupuncture combined with conventional drugs showed greater efficacy than conventional drugs alone in reducing TCMSS scores. Other therapies have not been sufficiently studied in relevant studies to provide meaningful comparisons with conventional drugs.

In terms of reducing CRP levels, acupuncture combined with conventional drugs was more effective than conventional drugs alone, while there was no statistically significant difference observed between blood-letting therapy and conventional drugs. Other therapies have been studied in too few relevant studies to allow for meaningful comparisons with conventional drugs.

In terms of reducing UA levels, acupuncture therapy, acupuncture therapy combined with conventional drugs, acupotomy combined with conventional drugs, blood-letting therapy, blood-letting therapy combined with conventional drugs, and fire acupuncture all demonstrated superior results compared to conventional drugs. And there was no statistically significant difference observed between acupotomy alone and conventional drugs.

In terms of reducing ESR (erythrocyte sedimentation rate), acupuncture combined with conventional drugs, blood-letting therapy, and blood-letting therapy combined with conventional drugs all showed greater efficacy compared to conventional drugs. Other therapies have not been sufficiently studied in relevant research to provide meaningful comparisons with conventional drugs.

Acupuncture, acupotomy, blood-letting therapy alone, and their combinations with conventional drugs, as well as electroacupuncture in conjunction with conventional medications, all showed superior efficacy in reducing VAS (Visual Analog Scale) scores compared to conventional drugs alone. Other therapies have not been sufficiently studied in relevant research to provide meaningful comparisons with conventional drugs.

#### 3.4.7 Examination of Heterogeneity

Within directly comparable meta-analyses, certain outcomes demonstrated variability. Examination of the original data suggested potential methodological heterogeneity in the included studies, primarily due to insufficient descriptions of blinding methods and allocation concealment. Additionally, clinical heterogeneity may have arisen from variations in patient populations, acupoint selections, and surgical techniques. However, because of insufficient detailed descriptions in the primary studies and a limited number of studies for specific outcomes, conducting subgroup analyses to explore the sources of heterogeneity was not possible.

### 3.5 Findings from Network Meta-Analysis

#### 3.5.1 Diagram of Evidence Network

A total of thirty-two studies ^9–20, 22–24, 26–31, 34–44^ contributed data on cure rates across 11 interventions, illustrating an incomplete closed-loop network. Seven studies^11, 14, 31, 36, 38, 39, 42^ reported TCMSS for 7 interventions without forming a closed loop. Eight articles^11, 28, 31–33, 36, 39, 43^ reported on CRP, involving eight intervention modalities, without _completing the closed loop. There were 27 articles 9-11, 13-15, 17-21, 23-25, 27-29, 31, 32, 34, 36, 38-40,_ ^42–44^ reporting on UA involving 11 interventions that did not form a closed loop. Nine articles^11, 13, 25, 28, 31, 32, 36, 39, 43^ reported on ESR involving 7 interventions without forming a closed loop. There were 17 studies^5, 10, 11, 16, 19, 23, 27–29, 32–36, 38, 40, 41^ reporting on VAS scores involving 9 interventions that did not form a closed loop. The width of the line connecting the two interventions correlates with the number of studies that compared them. The evidence network diagram for the cure rate is depicted in Figure 3.

**FIGURE 3.**
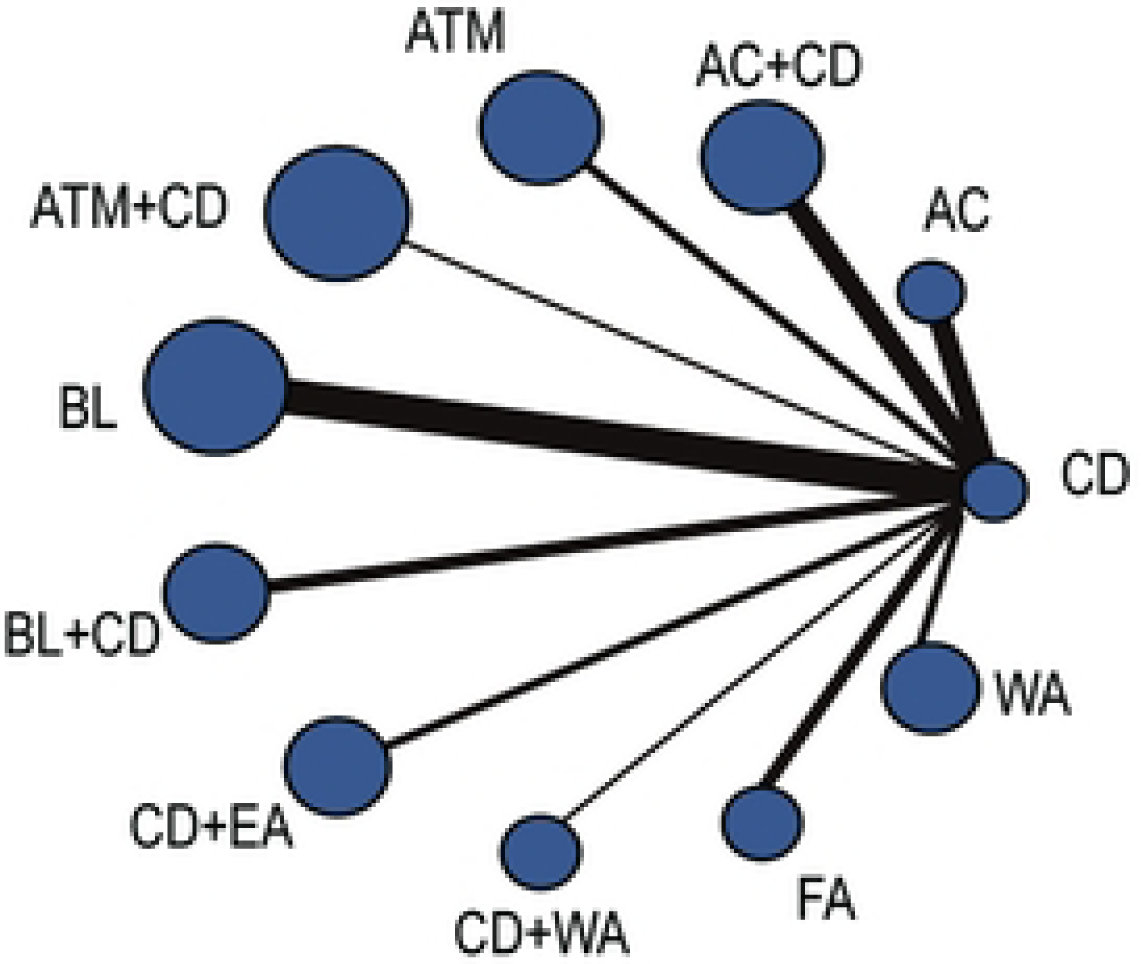
Evidence network diagram of the cure rate of differ.

#### 3.5.2 Results of the cure rate

Thirty-two studies ^9–20, 22–24, 26–31, 34–44^ reported cure rates. The results indicated that acupuncture [MD = 2.74 (95% CI: 1.49, 4.92)], acupuncture combined with conventional drugs [MD = 2.51 (95% CI: 1.35, 4.96)], blood-letting therapy [MD = 2.32 (95% CI: 1.48, 3.77)], and fire acupuncture [MD = 5.65 (95% CI: 2.31, 13.61)] demonstrated superior outcomes compared to conventional drugs. Additionally, fire acupuncture [MD = 4.09 (95% CI: 1.09, 14.75)] showed greater efficacy than electroacupuncture combined with conventional drugs. However, therapies such as acupuncture and acupuncture combined with conventional drugs did not show statistically significant differences compared to conventional drugs. Overall, no particularly significant differences in efficacy were observed among the various individual acupuncture treatments for the disease. (Table4) The probability ranking for improving cure rates was as follows: fire acupuncture > acupuncture > acupuncture combined with conventional drugs > blood-letting therapy > blood-letting therapy combined with conventional drugs > warm acupuncture therapy > warm acupuncture therapy combined with conventional drugs > electroacupuncture combined with conventional drugs > acupotomy combined with conventional drugs > acupotomy > conventional drugs.(Table5)

#### 3.5.3 Results of TCMSS

Seven studies^11, 14, 31, 36, 38, 39, 42^ reported TCMSS. The findings indicated no statistically significant difference in efficacy among the seven interventions: conventional drugs, acupuncture combined with conventional drugs, blood-letting therapy combined with conventional drugs, warm acupuncture therapy combined with conventional drugs, electroacupuncture combined with conventional drugs, fire acupuncture, and warm acupuncture therapy (Table 6). The probability ranking for reducing TCMSS was as follows: warm acupuncture therapy combined with conventional drugs > blood-letting therapy combined with conventional drugs > acupuncture combined with conventional drugs > fire acupuncture > electroacupuncture combined with conventional drugs > warm acupuncture therapy > conventional drugs(Table 7).

**Table 6.** NMA results of the TCMSS. (I Conventional drugs; 2 Acupuncture+ conventional drugs; 3 Blood-letting+ conventional drugs; 4 Warm acupuncture+ conventional drugs; *5* Fire acupuncture; 6 Warm acupuncture; 7 Electroacupuncture + conventional drugs.)

|  | X1 | X2 | X3 | X4 | X5 | X6 | X7 |
| --- | --- | --- | --- | --- | --- | --- | --- |
| 1 | 1 | -2.47<br>(-5.62,0.75) | -2.67<br>(-7.2,1.88) | -3.99<br>(-8.52,0.52) | -1.55<br>(-6.04,2.93) | -0.78<br>(-5.85,4.27) | -1.16<br>(-5.65,3.36) |
| 2 | 2.47<br>(-0.75,5.62) | 2 | -0.2<br>(-5.79,5.33) | -1.52<br>(-7.07,3.95) | 0.92<br>(-4.62,6.36) | 1.68<br>(-4.32,7.62) | 1.32<br>(-4.25,6.78) |
| 3 | 2.67<br>(-1.88,7.2) | 0.2<br>(-5.33,5.79) | 3 | -1.33<br>(-7.73,5.08) | 1.12<br>(-5.29,7.49) | 1.89<br>(-4.92,8.67) | 1.51<br>(-4.88,7.9) |
| 4 | 3.99<br>(-0.52,8.52) | 1.52<br>(-3.95,7.07) | 1.33<br>(-5.08,7.73) | 4 | 2.45<br>(-3.9,8.81) | 3.21<br>(-3.6,9.97) | 2.84<br>(-3.57,9.21) |
| 5 | 1.55<br>(-2.93,6.04) | -0.92<br>(-6.36,4.62) | -1.12<br>(-7.49,5.29) | -2.45<br>(-8.81,3.9) | 5 | 0.76<br>(-6,7.54) | 0.39<br>(-5.95,6.76) |
| 6 | 0.78<br>(-4.27,5.85) | -1.68<br>(-7.62,4.32) | -1.89<br>(-8.67,4.92) | -3.21<br>(-9.97,3.6) | -0.76<br>(-7.54,6) | 6 | -0.38<br>(-7.15,6.37) |
| 7 | 1.16<br>(-3.36,5.65) | -1.32<br>(-6.78,4.25) | -1.51<br>(-7.9,4.88) | -2.84<br>(-9.21,3.57) | -0.39<br>(-6.76,5.95) | 0.38<br>(-6.37,7.15) | 7 |

**Table 7.** Probability ranking results of the TCMSS. (1 Conventional drugs; 2 Acupuncture 18 + conventional drugs; 3 Blood-letting+ conventional drugs; 4 Warm acupuncture+ conventional drugs; *5* Fire acupuncture; 6 Warm acupuncture; 7 Elcctroacupuncture + conventional drugs.)

|  | V1 | V2 | V3 | V4 | V5 | V6 | V7 |
| --- | --- | --- | --- | --- | --- | --- | --- |
| 1 | 0.000 | 0.001 | 0.010 | 0.051 | 0.166 | 0.376 | 0.396 |
| 2 | 0.086 | 0.256 | 0.307 | 0.190 | 0.098 | 0.043 | 0.020 |
| 3 | 0.170 | 0.282 | 0.219 | 0.143 | 0.089 | 0.055 | 0.042 |
| 4 | 0.579 | 0.189 | 0.101 | 0.061 | 0.035 | 0.021 | 0.015 |
| 5 | 0.063 | 0.107 | 0.149 | 0.238 | 0.210 | 0.132 | 0.101 |
| 6 | 0.057 | 0.087 | 0.103 | 0.134 | 0.153 | 0.180 | 0.286 |
| 7 | 0.045 | 0.078 | 0.111 | 0.182 | 0.249 | 0.194 | 0.140 |

#### 3.5.4 Results of VAS Scores

Seventeen studies^5, 10, 11, 16, 19, 23, 27–29, 32–36, 38, 40, 41^ reported results of the VAS score. The results indicated that acupuncture combined with conventional drugs [MD = 1.75 (95% CI: 0.36, 3.14)], acupotomy [MD = 1.45 (95% CI: 0.45, 2.45)], and blood-letting therapy [MD = 1.17 (95% CI: 0.23, 2.11)] were more effective in reducing VAS scores compared to conventional drugs. Statistically significant differences were not observed in the remaining studies (Table 8). The ranking of the probability for reducing the VAS score was as follows: acupuncture combined with conventional drugs > acupotomy > fire acupuncture > blood-letting therapy > blood-letting therapy combined with conventional drugs > electroacupuncture combined with conventional drugs > acupuncture > warm acupuncture combined with conventional drugs > conventional drugs (Table 9).

**Table 8.**
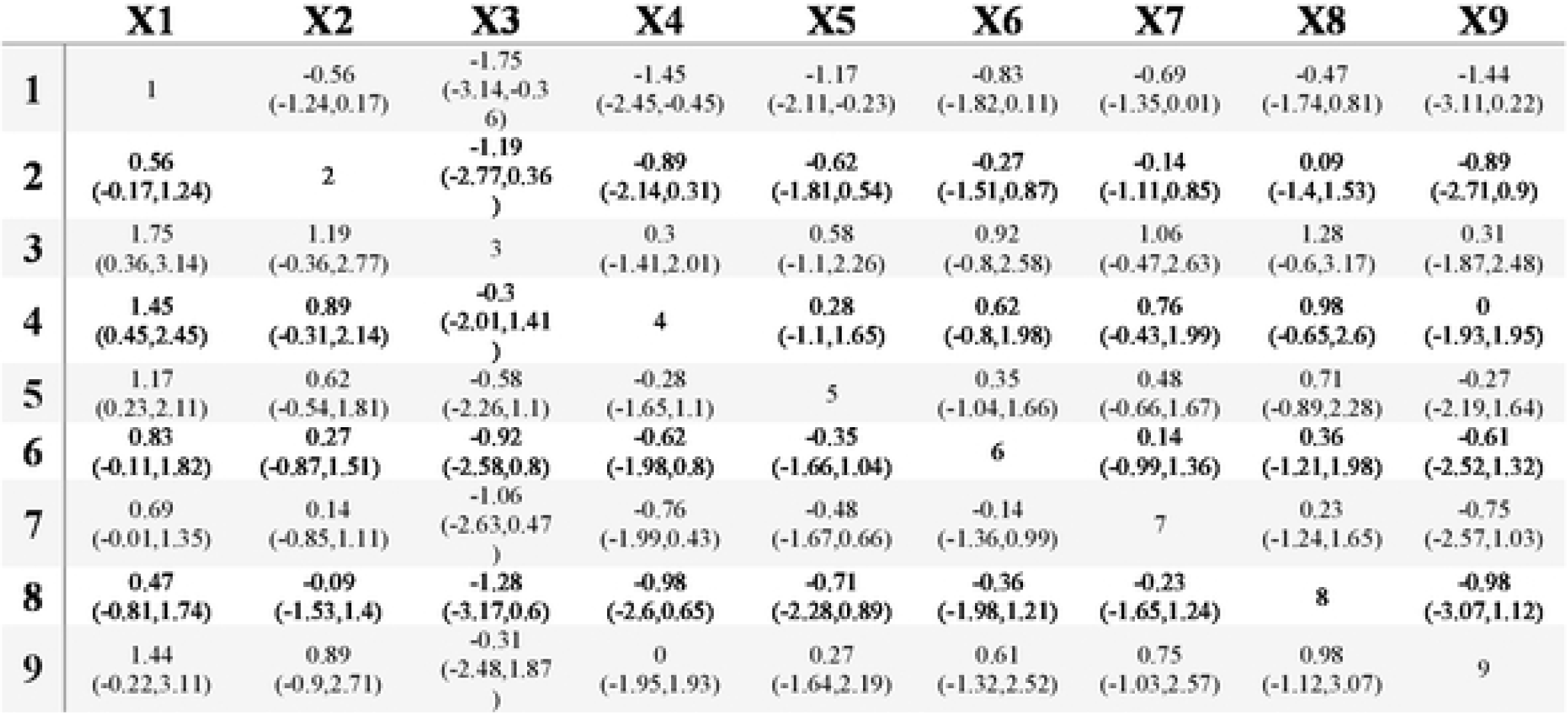
NMA results of the VAS score. (I Conventional drugs; 2 Acupuncture; 3 Acupuncture 21 + conventional drugs; 4 Acupotomy; *5* Blood-letting;6 Blood-letting+ conventional drugs; 7 Electroacupuncture + conventional drugs; 8 Wann acupuncture+ conventional drugs; 9 Fire acupuncture.)

**Table 9.** Probability ranking results of the VAS score. (I Conventional drugs; 2 Acupuncture; 3 Acupuncture+ conventional drugs; 4 Acupotomy; *5* Blood-letting; 6 Blood-letting + conventional drugs; 7 Electroacupuncture + conventional drugs; 8 Wann acupuncture+ conventional drugs; 9 Fire acupuncture.)

|  | V1 | V2 | V3 | V4 | V5 | V6 | V7 | V8 | V9 |
| --- | --- | --- | --- | --- | --- | --- | --- | --- | --- |
| 1 | 0.000 | 0.000 | 0.000 | 0.000 | 0.001 | 0.008 | 0.049 | 0.263 | 0.679 |
| 2 | 0.001 | 0.008 | 0.028 | 0.069 | 0.139 | 0.220 | 0.286 | 0.208 | 0.040 |
| 3 | 0.438 | 0.240 | 0.136 | 0.078 | 0.044 | 0.028 | 0.019 | 0.011 | 0.007 |
| 4 | 0.181 | 0.289 | 0.243 | 0.142 | 0.072 | 0.039 | 0.021 | 0.010 | 0.003 |
| 5 | 0.066 | 0.159 | 0.237 | 0.238 | 0.139 | 0.081 | 0.049 | 0.024 | 0.007 |
| 6 | 0.020 | 0.057 | 0.111 | 0.169 | 0.207 | 0.175 | 0.142 | 0.092 | 0.027 |
| 7 | 0.003 | 0.015 | 0.048 | 0.117 | 0.216 | 0.259 | 0.214 | 0.113 | 0.016 |
| 8 | 0.014 | 0.030 | 0.050 | 0.078 | 0.106 | 0.131 | 0.167 | 0.238 | 0.186 |
| 9 | 0.277 | 0.202 | 0.147 | 0.110 | 0.076 | 0.059 | 0.052 | 0.042 | 0.035 |

#### 3.5.5 Results of UA

_A total of twenty-seven studies9-11, 13-15, 17-21, 23-25, 27-29, 31, 32, 34, 36, 38-40, 42-44 presented results_ on UA. The results demonstrated that acupuncture [MD = 64.81 (95% CI: 13.75, 118.68)] and acupuncture combined with conventional drugs [MD = 77.85 (95% CI: 15.3, 151.89)] were more effective in reducing UA compared to conventional drugs. The differences observed in the remaining studies were not statistically significant (Table 10). The ranking of the probability of reducing UA was as follows: acupuncture combined with conventional drugs > acupotomy combined with conventional drugs > acupuncture > warm acupuncture therapy > blood-letting therapy > fire acupuncture > blood-letting combined with conventional drugs > acupotomy> warm acupuncture combined with conventional drugs > electroacupuncture combined with conventional drugs > conventional drugs(Table 11).

**Table 10.** NMA results of the UA. (I Conventional drugs; 2 Acupuncture; 3 Acupuncture+ conventional drugs; 4 Acupotomy; *5* Acupotomy + conventional drugs; 6 Blood-letting; 7 Blood-letting+ conventional drugs; 8 Wann acupuncture+ conventional drugs; 9 Fire acupuncture; 10 Wann acupuncture; and 11 Electroaeupuncture + conventional drugs.)

|  | X1 | X2 | X3 | X4 | X5 | X6 | X7 | X8 | X9 | X10 | X11 |
| --- | --- | --- | --- | --- | --- | --- | --- | --- | --- | --- | --- |
| 1 | 1 | -64.81<br>(-118.68,-<br>13.75) | -77.85<br>(-151.89,-<br>15.3) | -28.32(-12<br>0.69,61.76<br>) | -78.35<br>(-166.56,1<br>0.29) | -41.78(-93<br>.52,10.53) | -35.28<br>(-123.38,5<br>4.04) | -11.25<br>(-135.56,1<br>13.13) | -36.92<br>(-126.18,5<br>2.29) | -47.62<br>(-179.38,8<br>3.65) | -10.92<br>(-135.81,1<br>14.09) |
| 2 | 64.81<br>(13.75,11<br>8.68) | 2 | -13.07<br>(-102.61,6<br>8.07) | 36.34(-68.<br>69,141.91<br>) | -13.56<br>(-115.37,9<br>0.46) | 23.05(-49.<br>16,98.41) | 29.44<br>(-71.9,134<br>.22) | 53.47<br>(-80.33,19<br>0.92) | 27.79<br>(-74.1,132<br>.92) | 17.4<br>(-123.52,1<br>59.64) | 53.93<br>(-79.79,19<br>0.23) |
| 3 | 77.85<br>(15.3,151.<br>89) | 13.07<br>(-68.07,10<br>2.61) | 3 | 49.66(-59.<br>55,167.63<br>) | -0.53<br>(-106.33,1<br>17.47) | 36.13(-43.<br>96,127.61<br>) | 42.35<br>(-63.4,160<br>.26) | 66.32<br>(-69.21,21<br>4.35) | 41<br>(-65.7,159<br>.09) | 30.52<br>(-112.52,1<br>83.29) | 66.85<br>(-70.13,21<br>4.21) |
| 4 | 28.32<br>(-61.76,12<br>0.69) | -36.34<br>(-141.91,6<br>8.69) | -49.66<br>(-167.63,5<br>9.55) | 4 | -50.16<br>(-175.98,7<br>8.26) | -13.34(-11<br>7.02,92.87<br>) | -7.08<br>(-132.22,1<br>22.7) | 17.08<br>(-135.82,1<br>72.44) | -8.65<br>(-134.96,1<br>20.6) | -19.13<br>(-178.91,1<br>41.68) | 17.49<br>(-135.87,1<br>73.73) |
| 5 | 78.35<br>(-10.29,16<br>6.56) | 13.56<br>(-90.46,11<br>5.37) | 0.53<br>(-117.47,1<br>06.33) | 50.16(-78.<br>26,175.98<br>) | 5 | 36.74(-66.<br>16,139.55<br>) | 43.08<br>(-81.85,16<br>8.39) | 67.15<br>(-85.81,22<br>0.04) | 41.57(-83.<br>81,167.11<br>) | 30.97<br>(-127.3,18<br>9.5) | 67.7<br>(-86.02,21<br>9.82) |
| 6 | 41.78<br>(-10.53,93<br>.52) | -23.05<br>(-98.41,49<br>.16) | -36.13<br>(-127.61,4<br>3.96) | 13.34(-92.<br>87,117.02<br>) | -36.74(-13<br>9.55,66.16<br>) | 6 | 6.33<br>(-96.53,11<br>0.02) | 30.44<br>(-104.53,1<br>65.14) | 4.75<br>(-98.76,10<br>8.43) | -5.7<br>(-147.76,1<br>35.95) | 30.92<br>(-104.82,1<br>66.26) |
| 7 | 35.28<br>(-54.04,12<br>3.38) | -29.44<br>(-134.22,7<br>1.9) | -42.35<br>(-160.26,6<br>3.4) | 7.08(-122.<br>7,132.22) | -43.08(-16<br>8.39,81.85<br>) | -6.33<br>(-110.02,9<br>6.53) | 7 | 23.83<br>(-128.63,1<br>76.84) | -1.67<br>(-127.8,12<br>3.81) | -12.23<br>(-170.74,1<br>46.24) | 24.46<br>(-129.42,1<br>77.06) |
| 8 | 11.25<br>(-113.13,1<br>35.56) | -53.47<br>(-190.92,8<br>0.33) | -66.32<br>(-214.35,6<br>9.21) | -17.08(-17<br>2.44,135.8<br>2) | -67.15(-22<br>0.04,85.81<br>) | -30.44<br>(-165.14,1<br>04.53) | -23.83<br>(-176.84,1<br>28.63) | 8 | -25.57<br>(-178.18,1<br>27.64) | -36.09<br>(-217.25,1<br>45.37) | 0.41<br>(-176.42,1<br>76.39) |
| 9 | 36.92<br>(-52.29,12<br>6.18) | -27.79<br>(-132.92,7<br>4.1) | -41<br>(-159.09,6<br>5.7) | 8.65(-120.<br>6,134.96) | -41.57(-16<br>7.11,83.81<br>) | -4.75<br>(-108.43,9<br>8.76) | 1.67<br>(-123.81,1<br>27.8) | 25.57<br>(-127.64,1<br>78.18) | 9 | -10.78<br>(-169.87,1<br>48.53) | 25.91<br>(-128,179.<br>43) |
| 10 | 47.62<br>(-83.65,17<br>9.38) | -17.4<br>(-159.64,1<br>23.52) | -30.52<br>(-183.29,1<br>12.52) | 19.13(-14<br>1.68,178.9<br>1) | -30.97(-18<br>9.5,127.3) | 5.7<br>(-135.95,1<br>47.76) | 12.23<br>(-146.24,1<br>70.74) | 36.09<br>(-145.37,2<br>17.25) | 10.78<br>(-148.53,1<br>69.87) | 10 | 36.36<br>(-144.37,2<br>17.5) |
| 11 | 10.92<br>(-114.09,1<br>35.81) | -53.93<br>(-190.23,7<br>9.79) | -66.85<br>(-214.21,7<br>0.13) | -17.49(-17<br>3.73,135.8<br>7) | -67.7(-219<br>.82,86.02) | -30.92<br>(-166.26,1<br>04.82) | -24.46<br>(-177.06,1<br>29.42) | -0.41<br>(-176.39,1<br>76.42) | -25.91<br>(-179.43,1<br>28) | -36.36<br>(-217.5,14<br>4.37) | 11 |

**Table 11.**
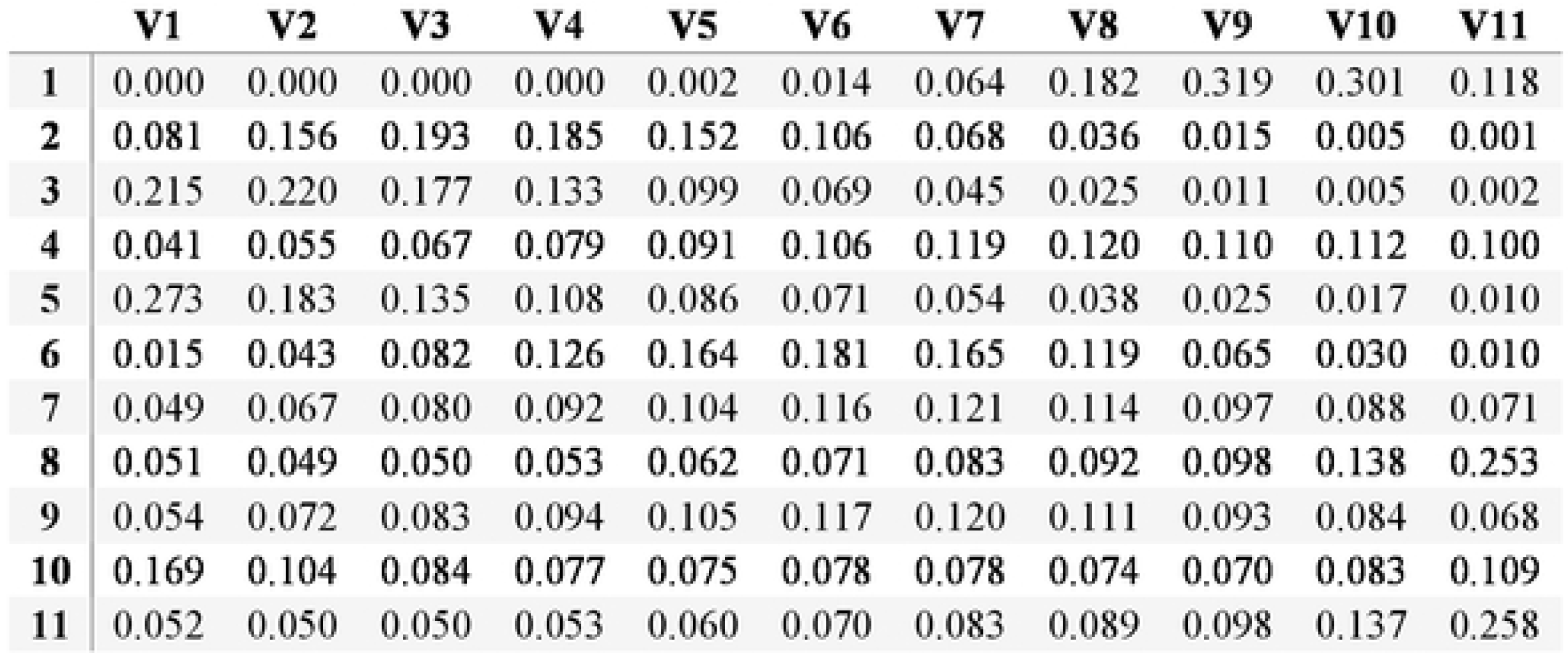
Probability ranking results of the UA. (I Conventional drugs; 2 Acupuncture; 3 Ac upunclure + conventional drugs; 4 Acupotomy; *5* AcupolOmy + conventional drugs; 6 Blood-letting; 7 Blood-letting+ conventional drugs; 8 Warm acupuncture+ conventional drugs; 9 Fire acupuncture; I0 Wann acupuncture; and 11 Eleclroacupunclure + conventional drugs.)

|  | V1 | V2 | V3 | V4 | V5 | V6 | V7 | V8 | V9 | V10 | V11 |
| --- | --- | --- | --- | --- | --- | --- | --- | --- | --- | --- | --- |
| 1 | 0.000 | 0.000 | 0.000 | 0.000 | 0.002 | 0.014 | 0.064 | 0.182 | 0.319 | 0.301 | 0.118 |
| 2 | 0.081 | 0.156 | 0.193 | 0.185 | 0.152 | 0.106 | 0.068 | 0.036 | 0.015 | 0.005 | 0.001 |
| 3 | 0.215 | 0.220 | 0.177 | 0.133 | 0.099 | 0.069 | 0.045 | 0.025 | 0.011 | 0.005 | 0.002 |
| 4 | 0.041 | 0.055 | 0.067 | 0.079 | 0.091 | 0.106 | 0.119 | 0.120 | 0.110 | 0.112 | 0.100 |
| 5 | 0.273 | 0.183 | 0.135 | 0.108 | 0.086 | 0.071 | 0.054 | 0.038 | 0.025 | 0.017 | 0.010 |
| 6 | 0.015 | 0.043 | 0.082 | 0.126 | 0.164 | 0.181 | 0.165 | 0.119 | 0.065 | 0.030 | 0.010 |
| 7 | 0.049 | 0.067 | 0.080 | 0.092 | 0.104 | 0.116 | 0.121 | 0.114 | 0.097 | 0.088 | 0.071 |
| 8 | 0.051 | 0.049 | 0.050 | 0.053 | 0.062 | 0.071 | 0.083 | 0.092 | 0.098 | 0.138 | 0.253 |
| 9 | 0.054 | 0.072 | 0.083 | 0.094 | 0.105 | 0.117 | 0.120 | 0.111 | 0.093 | 0.084 | 0.068 |
| 10 | 0.169 | 0.104 | 0.084 | 0.077 | 0.075 | 0.078 | 0.078 | 0.074 | 0.070 | 0.083 | 0.109 |
| 11 | 0.052 | 0.050 | 0.050 | 0.053 | 0.060 | 0.070 | 0.083 | 0.089 | 0.098 | 0.137 | 0.258 |

#### 3.5.6 Results of CRP

Eight studies^11, 28, 31–33, 36, 39, 43^ reported CRP results. The findings suggested no statistically significant differences among the interventions(Table 12). To assess the likelihood of CRP reduction, the outcomes were ranked as follows: warm acupuncture combined with conventional drugs > blood-letting therapy combined with conventional drugs > blood-letting therapy > acupuncture combined with conventional drugs > electroacupuncture combined with conventional drugs > conventional drugs > acupuncture(Table 13).

**Table 12.**
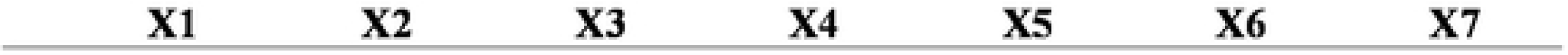
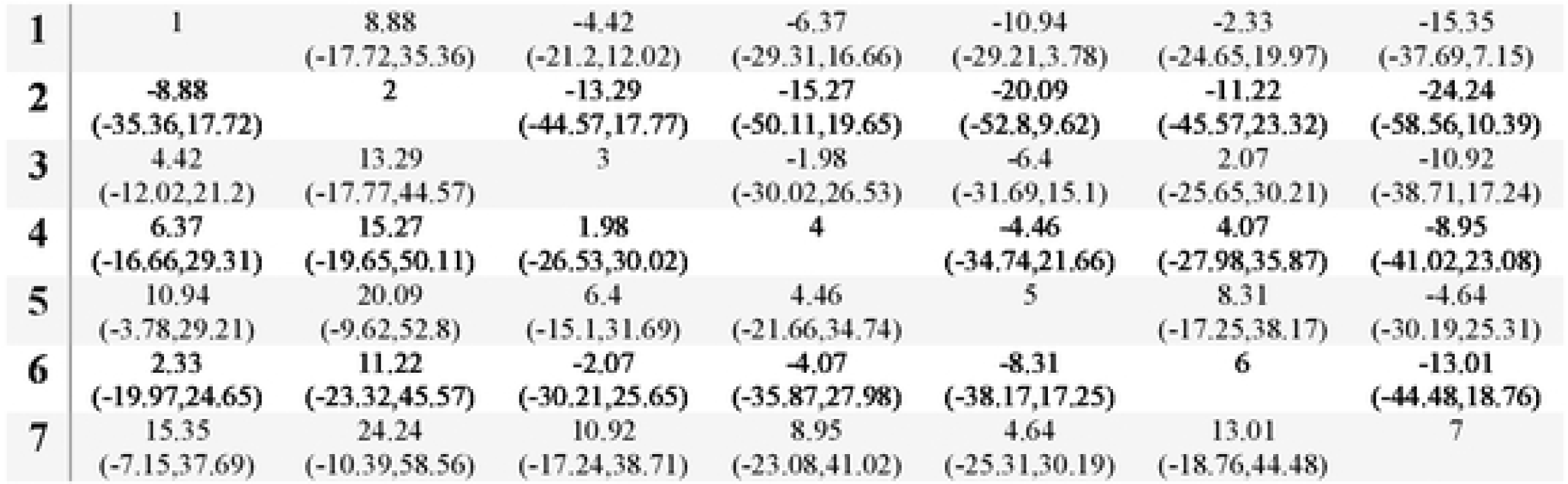
NMA results of the CRP. (I Conventional drugs; 2 Acupuncture; 3 Acupuncture+ conventional drugs; 4 Blood-letting; *5* Blood-letting+ conventional drugs; 6 Eleclroacupuncture + conventional drugs; 7 Wann acupuncture+ conventional drugs.)

| X1 | X2 | X3 | X4 | X5 | X6 | X7 |
| --- | --- | --- | --- | --- | --- | --- |

|  |  |  |  |  |  |  |  |
| --- | --- | --- | --- | --- | --- | --- | --- |
| 1 | 1 | 8.88 | -4.42 | -6.37 | -10.94 | -2.33 | -15.35 |
|  |  | (-17.72,35.36) | (-21.2,12.02) | (-29.31,16.66) | (-29.21,3.78) | (-24.65,19.97) | (-37.69,7.15) |
| 2 | -8.88 | 2 | -13.29 | -15.27 | -20.09 | -11.22 | -24.24 |
|  | (-35.36,17.72) |  | (-44.57,17.77) | (-50.11,19.65) | (-52.8,9.62) | (-45.57,23.32) | (-58.56,10.39) |
| 3 | 4.42 | 13.29 | 3 | -1.98 | -6.4 | 2.07 | -10.92 |
|  | (-12.02,21.2) | (-17.77,44.57) |  | (-30.02,26.53) | (-31.69,15.1) | (-25.65,30.21) | (-38.71,17.24) |
| 4 | 6.37 | 15.27 | 1.98 | 4 | -4.46 | 4.07 | -8.95 |
|  | (-16.66,29.31) | (-19.65,50.11) | (-26.53,30.02) |  | (-34.74,21.66) | (-27.98,35.87) | (-41.02,23.08) |
| 5 | 10.94 | 20.09 | 6.4 | 4.46 | 5 | 8.31 | -4.64 |
|  | (-3.78,29.21) | (-9.62,52.8) | (-15.1,31.69) | (-21.66,34.74) |  | (-17.25,38.17) | (-30.19,25.31) |
| 6 | 2.33 | 11.22 | -2.07 | -4.07 | -8.31 | 6 | -13.01 |
|  | (-19.97,24.65) | (-23.32,45.57) | (-30.21,25.65) | (-35.87,27.98) | (-38.17,17.25) |  | (-44.48,18.76) |
| 7 | 15.35 | 24.24 | 10.92 | 8.95 | 4.64 | 13.01 | 7 |
|  | (-7.15,37.69) | (-10.39,58.56) | (-17.24,38.71) | (-23.08,41.02) | (-25.31,30.19) | (-18.76,44.48) |  |

**Table 13.** Probability ranking results of the CRP. (I Conventional drugs; 2 Acupuncture; 3 Acupuncture+ conventional drugs; 4 Blood-letting; *5* Blood-letting+ conventional drugs; 6 Eleclroacupuncture + conventional drugs; 7 Wann acupuncture+ conventional drugs.)

|  | V1 | V2 | V3 | V4 | V5 | V6 | V7 |
| --- | --- | --- | --- | --- | --- | --- | --- |
| 1 | 0.000 | 0.005 | 0.037 | 0.151 | 0.337 | 0.394 | 0.076 |
| 2 | 0.019 | 0.033 | 0.047 | 0.060 | 0.072 | 0.119 | 0.651 |
| 3 | 0.045 | 0.113 | 0.215 | 0.267 | 0.176 | 0.128 | 0.056 |
| 4 | 0.112 | 0.189 | 0.229 | 0.173 | 0.118 | 0.111 | 0.068 |
| 5 | 0.216 | 0.379 | 0.226 | 0.102 | 0.044 | 0.023 | 0.009 |
| 6 | 0.053 | 0.085 | 0.137 | 0.186 | 0.218 | 0.198 | 0.123 |
| 7 | 0.555 | 0.196 | 0.109 | 0.061 | 0.035 | 0.028 | 0.016 |

#### 3.5.7 Results of ESR

Nine studies^11, 13, 25, 28, 31, 32, 36, 39, 43^ reported results for ESR. The results showed that acupuncture combined with conventional drugs [MD = 6.13 (95% CI: 0.55, 11.66)] and blood-letting therapy combined with conventional drugs [MD = 9.6 (95% CI: 3.06, 17.92)] demonstrated better outcomes compared to conventional drugs. The differences observed among the remaining interventions were not statistically significant (Table 14). For ranking the probability of reducing ESR, the results were as follows: blood-letting therapy combined with conventional drugs > blood-letting therapy alone > acupuncture alone > acupuncture combined with conventional drugs > warm acupuncture combined with conventional drugs > conventional drugs alone(Table15).

**Table 14.** NMA results of the ESR.(l. Conventional drugs; 2 Acupuncture; 3 Acupuncture+ conventional drugs; 4 Blood-letting; 5 Blood-lelling + conventional drugs; 6 Wann acupuncture+ conventional drugs.)

|  | X1 | X2 | X3 | X4 | X5 | X6 |
| --- | --- | --- | --- | --- | --- | --- |
| 1 | 1 | -6.04 | -6.13 | -7.84 | -9.6 | -4.55(-19.14,10.02) |
|  |  | (-17.07,5.18) | (-11.66,-0.55) | (-14.11,1.37) | (-17.92,-3.06) |  |
| 2 | 6.04 | 2 | -0.08 | -1.7 | -3.61 | 1.43(-16.86,19.66) |
|  | (-5.18,17.07) |  | (-12.63,12.24) | (-14.12,12.9) | (-17.86,8.95) |  |
| 3 | 6.13 | 0.08 | 3 | -1.73 | -3.46 | 1.59(-13.85,17.04) |
|  | (0.55,11.66) | (-12.24,12.63) |  | (-9.78,9.32) | (-13.71,4.82) |  |
| 4 | 7.84 | 1.7 | 1.73 | 4 | -1.78 | 3.09 |
|  | (-1.37,14.11) | (-12.9,14.12) | (-9.32,9.78) |  | (-15,6.84) | (-14.02,18.61) |
| 5 | 9.6 | 3.61 | 3.46 | 1.78 | 5 | 5.14 |
|  | (3.06,17.92) | (-8.95,17.86) | (-4.82,13.71) | (-6.84,15) |  | (-10.63,21.88) |
| 6 | 4.55 | -1.43 | -1.59 | -3.09 | -5.14 | 6 |
|  | (-10.02,19.14) | (-19.66,16.86) | (-17.04,13.85) | (-18.61,14.02) | (-21.88,10.63) |  |

**Table 15.**
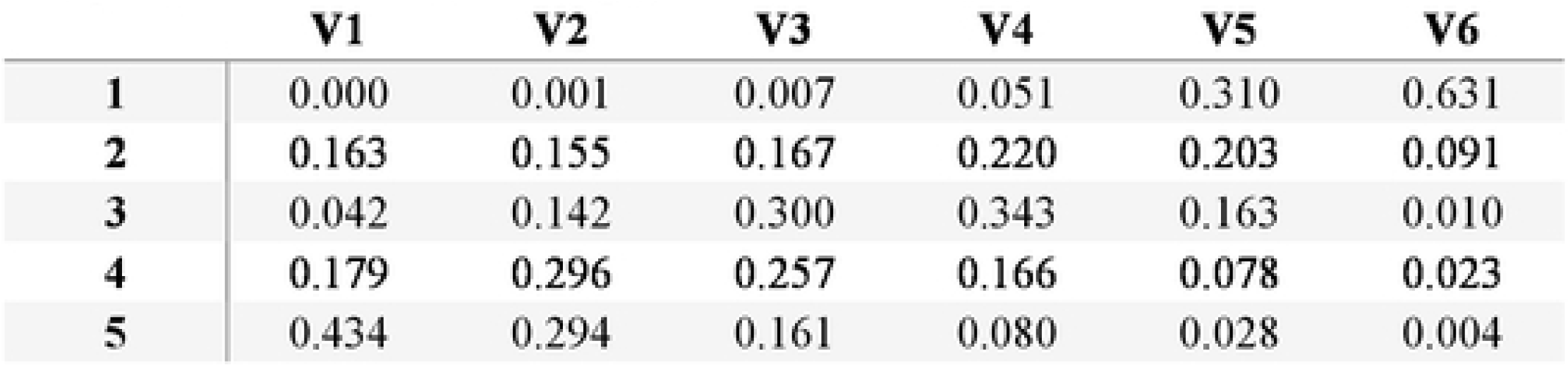
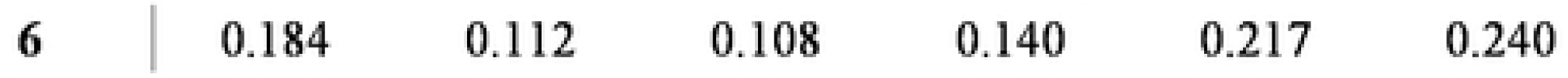
Probability ranking results of the ESR. (I Conventional drugs; 2 Acupuncture; 3 Acupuncture + conventional drugs; 4 Blood-lclting ; *5* Blood-letting + conventional drugs; 6 Wann acupuncture+ conventional drugs.)

#### 3.5.7 Evaluation of Small Sample Effects

A comparison-adjusted funnel plot depicting the cure rate for primary outcome indicators was created using R version 4.4.0 software. Combined with Figure 4 and the Egger test of P>0.05, this suggests that potential publication bias or small-sample effects within the research network are highly unlikely.

**FIGURE 4.**
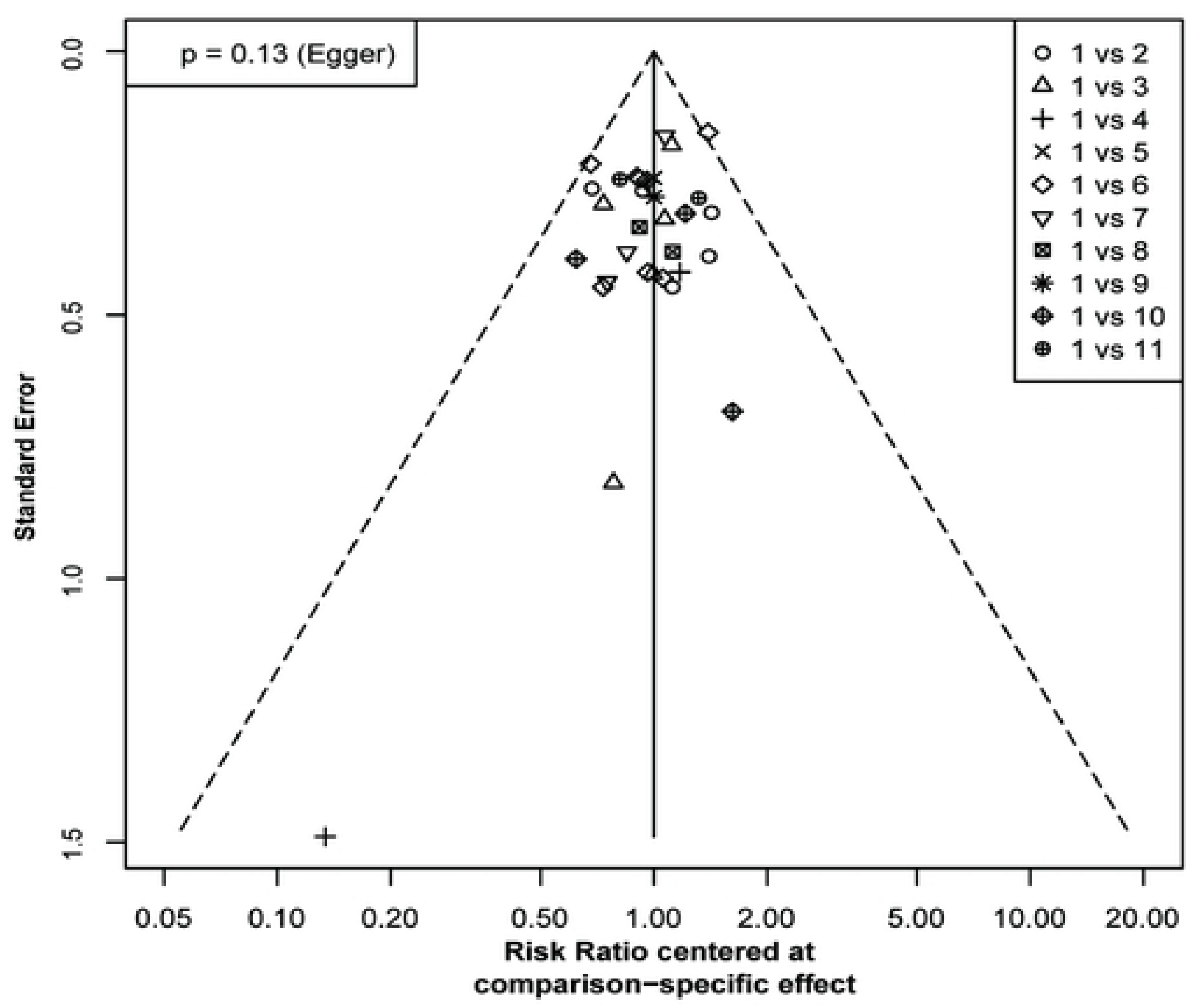
Comparison-correction funnel plot of the cure rate.

### 3.6 Adverse Reactions

Adverse reactions were reported in 10 studies^10, 11, 16, 22, 25, 27, 28, 32, 33, 38^ (10, 11, 16, 22, 25, 27, 28, 32, 33, 38)(Table 16). Overall, the adverse reactions associated with various acupuncture-related therapies were generally lower than those observed in the conventional drug group, and no serious adverse reactions related to acupuncture were reported.

**Table 16.**
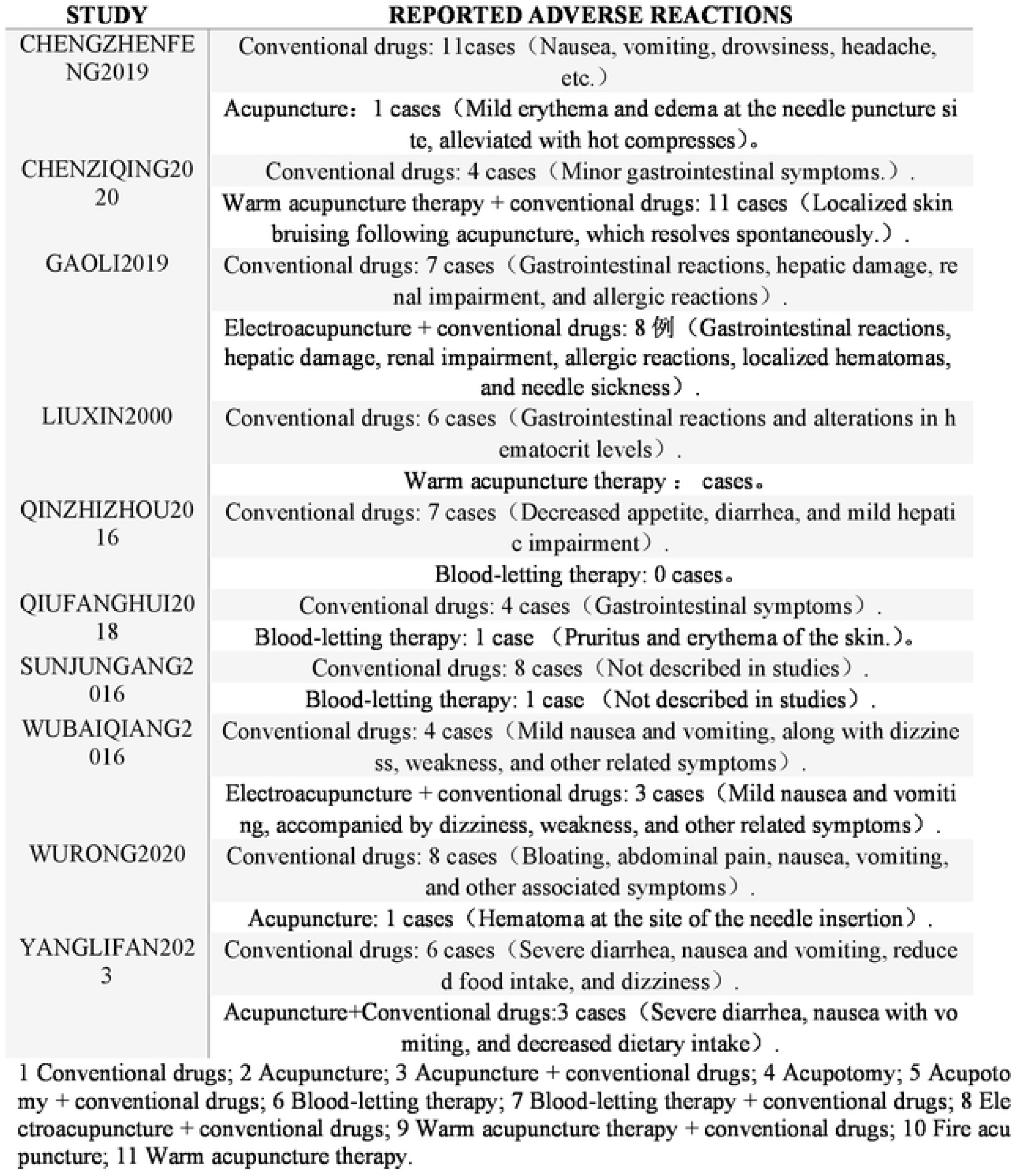
Adverse reaction of the included studies.

Table 16 Adverse reaction of the included studies.

## 4 Discussion

Acute gouty arthritis (AGA) is characterized by the deposition of monosodium urate crystals within joints, initiating an inflammatory cascade that culminates in the release of various inflammatory mediators into the joint cavity^8^. This process manifests clinically as localized redness, swelling, warmth, and pain in the affected joints. As the most prevalent inflammatory arthritis in men, gout affects 1-2 percent of adults in developed countries^45–47^. Gouty arthritis resulting from hyperuricemia is significantly linked to an increased risk of hypertension, diabetes, renal disease, and cardiovascular disease^48^. Therefore, early intervention in gouty arthritis will significantly improve patient prognosis. ^1, 49^. Currently, the primary first-line therapeutic agents in clinical practice are colchicine and non-steroidal anti-inflammatory drugs (NSAIDs), supplemented by a short glucocorticoid therapy if needed for anti-inflammatory purposes. Research indicates that drugs like colchicine, non-steroidal anti-inflammatory drugs (NSAIDs), and glucocorticoids can lead to significant side effects and may induce resistance in certain patients, thereby potentially diminishing treatment efficacy^50^. Acupuncture serves as a pivotal component in the management of acute gouty arthritis, offering a dependable and safe alternative therapy ^49^. Research has shown the effectiveness and safety of acupuncture when combined with conventional medications^14, 17, 21, 25^. However, current research emphasizes the importance of identifying the optimal combination of therapies^51^.

This study aimed to evaluate the impact of acupuncture-related therapies combined with conventional drugs on outcomes such as the cure rate, VAS (Visual Analog Scale), TCMSS (Traditional Chinese Medicine Symptom Score), UA (uric acid levels), CRP (C-reactive protein), and ESR (erythrocyte sedimentation rate) in patients diagnosed with acute gouty arthritis (AGA). The Visual Analogue Scale (VAS) is widely employed in assessing remission in gout patients by evaluating pain levels and continuously monitoring disease activity. Serological biomarkers such as CRP, ESR, and UA are crucial for evaluating the active stage of gout, reflecting the extent of inflammation and tissue damage in those affected. The study results indicated that fire acupuncture was the most effective treatment in terms of significantly increasing the cure rate. The combination of acupuncture with conventional drugs demonstrated significant efficacy in reducing VAS scores. Moreover, this combined approach has been identified as highly effective in reducing UA levels. Blood-letting therapy combined with conventional drugs was found to be the most effective in reducing ESR. However, there were no statistically significant differences observed among the various interventions in terms of reducing TCMSS and CRP.

Based on the study findings, the highest rankings were achieved by: blood-letting combined with conventional drugs, acupuncture combined with conventional drugs, and fire acupuncture. Recent research indicates that acupuncture is effective in reducing pain and improving the quality of life in patients with AGA. Its efficacy may be associated with the modulation of inflammatory factors, serum metabolites, and neurotransmitter pathways. Both blood-letting and fire acupuncture techniques have evolved from and refined traditional acupuncture theory. Blood-letting therapy involves the therapeutic use of acupuncture or knives to puncture or incise specific points and areas of the body, facilitating the controlled release of small amounts of blood. Blood-letting therapy has been shown to modulate the immune response by decreasing levels of IL-1, IL-8, and TNF-α^52–54^, while simultaneously upregulating levels of IL-4 and IL-10^55, 56^, and influencing the TLR4/IL-1β signaling pathway. Simultaneously, blood-letting therapy can reduce serum metabolite levels of methionine, accelerate uric acid excretion, influence the L-arginine/nitric oxide signaling pathway in the human body, regulate neurotransmitters, and promote the alleviation of symptoms in patients with acute gouty arthritis^25^. Fire acupuncture is a therapeutic technique in which acupuncture, heated red-hot over a fire, is rapidly inserted into the body to treat various diseases. Fire acupuncture can effectively halt the onset and progression of acute gouty joint inflammation by inhibiting the expression and activation of the NALP3 inflammasome and IL-1β inflammatory signaling pathways. Interestingly, in a rat model of acute gouty arthritis, fire acupuncture has been observed to rapidly reduce serum levels of NALP3 and IL-1β, similar to the effects of colchicine. Furthermore, fire acupuncture has been shown to ameliorate pathological changes in the synovial membrane of joints to a degree comparable to that of colchicine^57,58^.

Nevertheless, this study has some limitations. One major issue is that several of the included studies did not clearly detail their methods for randomization, allocation concealment, or blinding, potentially affecting the validity and reliability of the results. Secondly, the small sample sizes in the included studies may have limited the precision and generalizability of the results. Thirdly, variability existed among the included studies regarding the type and dosage of conventional drugs, as well as the selection of acupuncture points, depth, and duration of acupuncture-related therapies. This variability may have contributed to clinical heterogeneity across the studies. Fourthly, the limited number of primary studies and the omission of additional acupuncture methods such as bloodletting therapy and acupoint injections prevented a comprehensive evaluation of the effectiveness of all acupuncture-related interventions.

Overall, a comprehensive comparison of outcome metrics across the six different therapies demonstrated that fire acupuncture was the most effective treatment in terms of significantly increasing the cure rate. Combining acupuncture with conventional drugs was found to be highly effective in reducing VAS scores and lowering UA levels. Acupuncture combined with conventional drugs has shown to be the most effective approach for lowering UA levels. Blood-letting therapy combined with conventional drugs was found to be the most effective in reducing ESR. In clinical practice, treatments should be selected based on individual circumstances and clinical presentation. Given the limited number of available studies, many of which are of suboptimal quality, there is a need for additional multicenter, large-scale, prospective randomized controlled trials (RCTs) to confirm these results.

## AUTHOR CONTRIBUTIONS

Yifan Liuconceptualized the study and wrote the manuscript.. Yuxiao Cai analyzed the data and wrote the manuscript. Qinglong Kang; Shiji Zhu; Qianqian Wang and Chunmei Sun collected the data. Yin Gu revised the manuscript.

## ADDITIONAL INFORMATION

There are no declared conflicts of interest.

## DATA AVAILABILITY

Registration of Systematic Review: This study has been registered with PROSPERO under registration number CRD42021278233. Detailed information can be accessed at PROSPERO.

## ACKNOWLEDGMENTS

We thank the subjects for their committed participation in this study.

